# Impact of a sanitation intervention on quality of life and mental wellbeing in low-income urban neighbourhoods of Maputo, Mozambique

**DOI:** 10.1101/2022.02.25.22271508

**Authors:** Ian Ross, Giulia Greco, Zaida Adriano, Rassul Nala, Joe Brown, Charles Opondo, Oliver Cumming

**Affiliations:** Department of Global Health and Development, London School of Hygiene and Tropical Medicine, UK; Department of Disease Control, London School of Hygiene & Tropical Medicine, UK; WE Consult, Maputo, Mozambique; Instituto Nacional de Saúde, Maputo, Mozambique; Department of Environmental Sciences and Engineering, University of North Carolina, Chapel Hill, USA; Department of Medical Statistics, London School of Hygiene and Tropical Medicine, UK; Nuffield Department of Population Health, University of Oxford, UK

## Abstract

**Objectives:** Toilet users often report valuing privacy and safety more highly than reduced disease, but effects of urban sanitation interventions on such outcomes have never been assessed quantitatively. In this study, we evaluate the impact of a shared sanitation intervention on quality of life (QoL) and mental wellbeing.

**Design:** We interviewed individuals living in intervention and control clusters of a recent non-randomised controlled trial, and used generalised linear mixed regression models to make an observational comparison of outcomes.

**Setting:** low-income unsewered areas of Maputo City, Mozambique

**Participants:** We interviewed 424 participants, 222 from the prior trial’s intervention group, and 202 from the control group.

**Interventions:** The control group used low-quality pit latrines. The intervention group received high-quality shared toilets, contributing 10-15% of capital cost.

**Outcomes:** Our primary outcome was the sanitation-related quality of life (SanQoL) index, which applies respondent-derived weights to combine toilet users’ perceptions of sanitation-related disgust, privacy, safety, health, and shame. Secondary outcomes were the WHO-5 mental wellbeing index and a sanitation visual analogue scale.

**Results:** The intervention group experienced a 1.6 standard deviation gain in SanQoL compared to the control group. This adjusted SanQoL gain was 0.34 (95% CI: 0.29-0.38) on a 0-1 scale with control mean 0.49. Effect sizes were largest for safety and privacy attributes. Intervention respondents also experienced a 0.2 standard deviation gain in mental wellbeing. The adjusted gain was 6.2 (95% CI: 0.3-12.2) on a 0-100 scale with control mean 54.4.

**Conclusions:** Quality of life outcomes highly valued by toilet users and can be improved by sanitation interventions. Such outcomes should be measured in future sanitation trials, to help identify interventions which most improve people’s lives. Since SanQoL weights are derived from respondent valuation, the measure can also be used in economic evaluation.

## 1. Introduction

Nearly two billion people globally lack access to “basic” sanitation.^1^ This deficit leads to 400,000 deaths from diarrhoeal disease annually, as well to the helminth infections and other diseases.^2^ However, inadequate sanitation has further negative consequences for health broadly defined,^3^ including poorer outcomes across various domains of quality of life (QoL) such as privacy, safety, and dignity.^4–6^ For example, women and girls are at risk of infringements of safety and privacy, including violence, when using inadequate sanitation or practising open defecation.^5,7,8^ Smelling faeces and touching surfaces in low-quality latrines can trigger disgust and shame.^9–11^

Sanitation interventions can have positive health externalities,^12,13^ and an excreta-free environment is a public good in that it is non-rival and non-excludable.^14^ Provision of this public good is supported by household investments in toilets, which are private goods.^15,16^ In studies of what toilet users most value about sanitation, QoL outcomes such as privacy, safety, and dignity are usually high up their list, and often above disease prevention.^9,17,18^ Therefore, the expected QoL payoff from a private sanitation investment is an important determinant of whether the public good of an excreta-free environment is achieved.^15,16^ Each sanitation-related QoL outcome (such as privacy, safety, and dignity) maps onto one of seven “objective features of QoL” identified by a recent commission.^19^ Sanitation-related QoL, then, is the subset of overall QoL which is directly affected by sanitation practices or services.^10,20^

Different sanitation interventions plausibly improve sanitation-related QoL to different extents, but most sanitation impact evaluations focus on infectious diseases.^21^ A systematic review of the relationship between sanitation and mental and social well-being concluded that innovation in quantitative measurement of such outcomes is a research priority.^5^ One quantitative study has explored the association in the general population between urban sanitation access and mental wellbeing outcomes,^22^ and another has done so in rural areas.^23^ However, no studies have quantified the broader QoL effect of a specific urban sanitation intervention, with one such study ongoing.^24^ In this study, we evaluate the effect of a shared urban sanitation intervention on sanitation-related QoL and mental wellbeing in urban Mozambique, as compared to existing use of shared pit latrines.

## 2. Methods

### Setting

Maputo City, Mozambique’s capital, has a population of 1.1 million, with the majority living in basic settlements with unpaved roads.^25^ Pit latrines are used by 41% of people, and less than half of faecal waste is safely managed.^26^ Our study site comprises low-income neighbourhoods in a 10km^2^ area of the Nhlamankulu district (detail and maps in Online Appendix A). In this area, the poorest people live in informally-walled ‘compounds’ with many households sharing the same toilet. Low-quality pit latrines are common, with unlined pits and squatting slabs made of wood or tyres, and no water seal (u-bend) providing a barrier to smells and flies. Latrine walls are usually made with scrap corrugated iron or plastic sheeting, doors are makeshift, and roofs are uncommon.

### Study design

We report an observational study sampling households from the intervention and control clusters of a prior non-randomised trial with a controlled before-and-after design (clinicaltrials.gov, NCT02362932).^27^ Intervention compounds in the Maputo Sanitation trial (MapSan) were identified in 2015-16 using the following criteria: (i) inhabitants sharing poor-quality sanitation; (ii) at least 12 inhabitants; (iii) inhabitants willing to contribute financially to construction; (iv) legal on-plot piped water; (v) located in pre-defined neighbourhoods; (vi) sufficient space for construction; (vii) accessible for transportation of construction materials; (viii) water table low enough for septic tank installation.

The MapSan trial enrolled the compound if there was at least one resident child younger than 48 months. As each intervention compound was enrolled, investigators concurrently enrolled a control compound according to criteria 1-4 above and by number of inhabitants (cluster size). Control compounds were located in the same or adjacent neighbourhoods. MapSan concluded that the intervention had no effect on any measure of child health, with a 24-month diarrhoea prevalence ratio of 0.84 (95% CI: 0.47 – 1.51).^7^

### Participants

Eligible participants for our study were people aged 18 or over and: (i) living in MapSan intervention or control compounds for four or more years, since before the intervention; (ii) still using the type of toilet consistent with intervention/control status (e.g. pit latrine if control). The first criterion ensured that, prior to the intervention, all our participants had been using a pit latrine without a water seal in that same compound they still lived on. This aimed to reduce risk of selection bias, because there had been migration out of and into MapSan-enrolled compounds since 2015. The second criterion was motivated by the knowledge that a small number of control compounds had: (a) received NGO sanitation interventions under a post-MapSan programme; or (b) autonomously upgraded their toilets.

We aimed to recruit two people per compound (one man, one woman) from different households. We used trial records to pre-assess eligibility for the 593 MapSan compounds (clusters), leading to the exclusion of 35 (Figure 1). The two lists of remaining MapSan intervention and control compounds were then randomly reordered, and visited in that new order. Procedures for sampling individuals within a compound are in Online Appendix B, but are summarised as listing of eligible individuals followed by random sampling. A team of four fieldworkers interviewed participants in Portuguese, unless the participant preferred to talk in Changana, a local language in which all interviewers were fluent.

**Figure 1:**
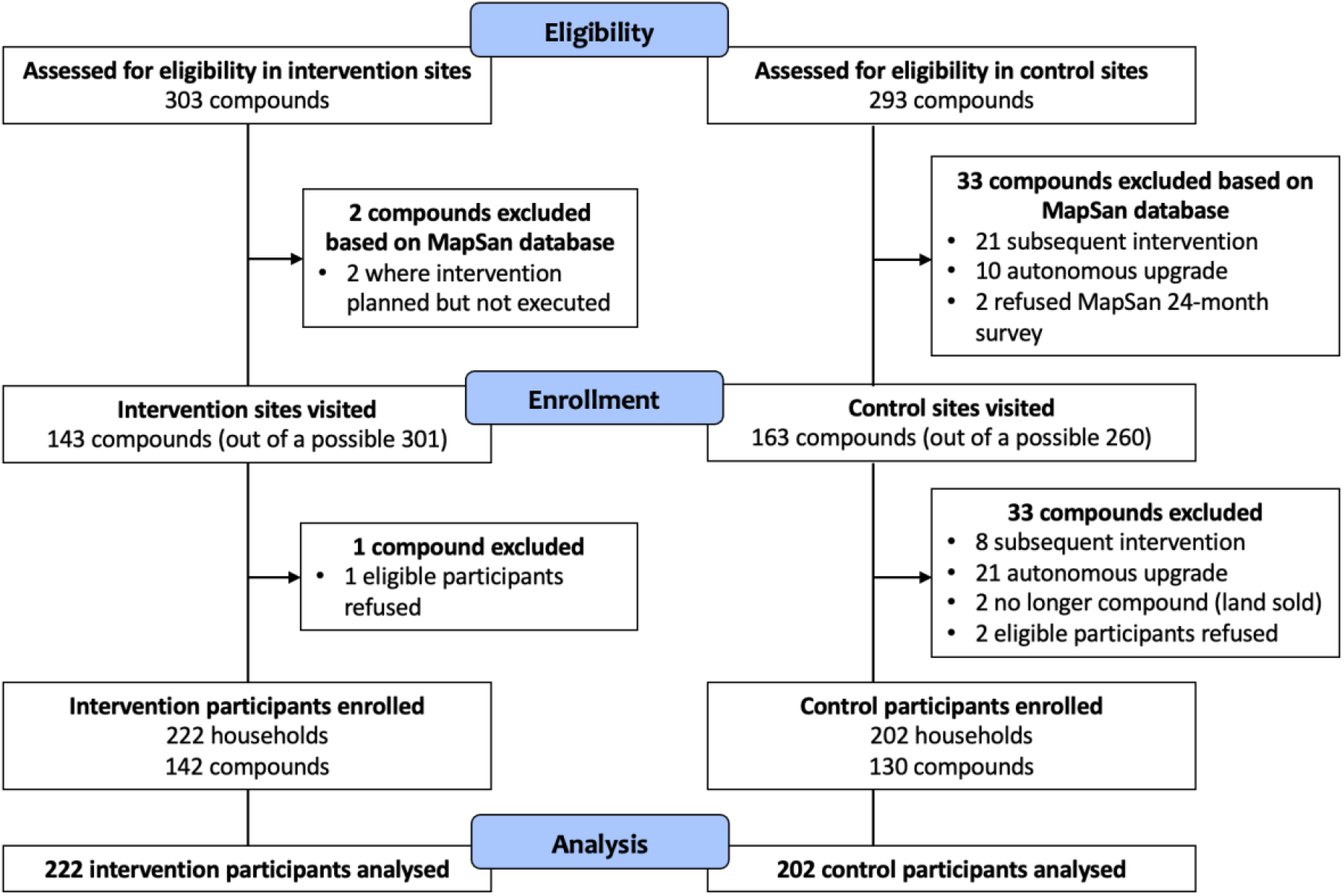
Participant flow diagram showing eligibility, enrolment and analysis

### Interventions

The intervention we evaluated was implemented by Water & Sanitation for the Urban Poor (WSUP), a non-government organisation. Compounds were provided with a subsidised pour-flush toilet with a water seal and concrete superstructure, discharging to a septic tank with soakaway (detail/photos in Online Appendix A). Two toilet types were provided, depending on user numbers. A shared toilet (ST) with one stance (cubicle) was designed for around 15 people, while a Community Sanitation Block (CSB) with two stances was designed for at least 21 people. Both STs and CSBs had metal doors lockable from the inside. Compound inhabitants had to pay a 10-15% capital contribution – approximately US$ 120 for CSB (2015 prices) and US$ 80 for ST.

### Outcomes

The primary outcome is an index of sanitation-related quality of life (SanQoL), deriving from a capability-based questionnaire informed by qualitative research.^10,20^ SanQoL measures aspects of self-perceived quality of life which are directly affected by sanitation practices or services. Validity and reliability of SanQoL were previously assessed in the Maputo setting through cognitive interviews and psychometric analysis.^20^ The five SanQoL attributes are disgust, health, privacy, safety, and shame, measured on a four-level frequency scale (Table 1). Responses are combined as an index by weighting attributes according to their relative importance, assessed via a ranking exercise undertaken with all study participants (Online Appendix B). The ensuing weights, which sum to 1 (Table 1), were used to calculate SanQoL index values on a 0-1 scale. Higher scores are better, with zero representing ‘no sanitation-related capability’ and one ‘full sanitation-related capability’. Histograms of outcome variables by group are in Online Appendix C.

**Table 1:** SanQoL attributes and weights

| Attribute | Psychometric item | Responses | Weight in index valuation |
| --- | --- | --- | --- |
| **Disgust** | Can you use the toilet without feeling disgusted? | Always<br>Sometimes<br>Rarely<br>Never | 0.22 |
| **Health** | Can you use the toilet without worrying that it spreads diseases? |  | 0.29 |
| **Privacy** | Can you use the toilet in private, without being seen? |  | 0.20 |
| **Shame** | Can you use the toilet without feeling ashamed for any reason? |  | 0.13 |
| **Safety** | Are you able to feel safe while using the toilet? |  | 0.16 |

In estimating index values, attribute-level scores are applied as “always”=3, “never”=0, etc. (formulae in Online Appendix B).

The second outcome is a sanitation visual analogue scale (VAS). We asked people to indicate on a paper-based 0-10 scale how they felt about their “level of sanitation today”, where zero is “worst imaginable sanitation” and ten is “best imaginable sanitation” (Online Appendix B, Figure B-2). The rationale for including the sanitation VAS was to explore whether an effect size comparable to that for SanQoL would be seen when people rated their level of sanitation directly rather than via questionnaire items.

The third outcome is the WHO-5 mental wellbeing index, a multi-attribute instrument for assessing subjective mental wellbeing.^28^ It comprises five items related to feeling cheerful, calm, active, well-rested, and finding enjoyment in daily life, scored on a frequency scale (Online Appendix B). Scores are summed with equal weighting and rescaled to 0-100, with higher scores better. The rationale for including WHO-5 was that mental wellbeing is thought to be influenced by sanitation but, unlike SanQoL, is not specific to sanitation.^5^

### Statistical analyses

The sample size calculation was estimated according to a formula for the comparison of two means with 80% power and significance at 0.05. The required sample size to detect a 0.05 mean difference in SanQoL with a standard deviation of 0.15 and intracluster correlation coefficient (ICC) of 0.4 was estimated as 398. We computed a wealth index using principal components analysis,^29^ using the asset list from the most recent Mozambican Demographic and Health Survey.

We analysed participants according to trial arm, to test the overarching hypothesis that the intervention was associated with an improvement in quality of life. Specific hypotheses and associated rationales are in Online Appendix B but in summary: (i) hypothesis 1a – the intervention is associated with higher SanQoL (with 1b and 1c the same for VAS and WHO-5); (ii) hypothesis 2a – across all three outcomes, effects are larger for women than men (with 2b the same but for larger effects on people aged over-60 than under-60); (iii) hypothesis 3 – some SanQoL attributes contribute more to overall differences than others. P-values less than 0.05 were considered statistically significant evidence of association, and we ran analyses in Stata 16.

To test hypothesis 1a, we used a generalised linear mixed model (GLMM), with gaussian distribution and identity link.

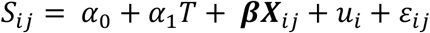

where:

*S*_*ij*_ represents the SanQoL index value for individual *j* in compound *i*

*T* is 1 for intervention and 0 for control

***X***_*ij*_ is a vector of covariates

*α*_0_ is a constant with no interpretation in this case

*α*_1_ is a coefficient and ***β*** a vector of coefficients

*u*_*i*_ is a random effect at the compound level

*ε*_*ij*_ is the error term

Standard errors were clustered at the compound level, since the intervention was delivered at this level, requiring the assumption that errors are not correlated across compounds. Spatial distribution of compounds was within one small area of Maputo (map in Online Appendix A). We included two types of covariates in ***X***_*ij*_. First, we adjusted for characteristics which were unbalanced at the 5% level between groups (Table 2), i.e. the wealth index only. Second, we included binary variables for gender and being elderly (aged over-60), since these were hypothesised to affect the association between exposure and outcome (Online Appendix B). Only 2 participants had missing data for outcomes or covariates (one for WHO-5, one for the wealth index).

**Table 2:** Characteristics of sample

|  | Control<br>(n=202) | Intervention<br>(n=222) | P-value for<br>difference<br>(t-test) |
| --- | --- | --- | --- |
| **Demographic characteristics** |  |  |  |
| Respondent is male | 101 (50%) | 103 (46%) | 0.459 |
| Respondent age | 38.4 (14.9) | 41.2 (15.6) | 0.059* |
| Respondent has a partner | 107 (53%) | 107 (48%) | 0.327 |
| Household size | 5.0 (2.8) | 5.2 (3.2) | 0.405 |
| Number of children under-14 | 1.4 (1.5) | 1.2 (1.6) | 0.122 |
| **Wealth index** |  |  |  |
| Wealth index score | -0.13 (1.00) | 0.12 (0.99) | 0.010** |
| *Dwelling has cement or tiled floor* | 184 (91%) | 210 (95%) | 0.160 |
| *Dwelling has concrete exterior walls* | 140 (69%) | 143 (64%) | 0.287 |
| *Access to electricity connection* | 167 (83%) | 192 (86%) | 0.277 |
| *Access to piped water connection* | 199 (99%) | 217 (98%) | 0.563 |
| *Household cooks indoors* | 114 (56%) | 114 (51%) | 0.295 |
| *Household owns television* | 153 (76%) | 184 (83%) | 0.069* |
| *Household owns fridge* | 98 (49%) | 128 (58%) | 0.060* |
| *Household owns mobile phone* | 166 (82%) | 191 (86%) | 0.278 |
| *Household owns bicycle* | 7 (3%) | 6 (3%) | 0.656 |
| *Household owns radio* | 63 (31%) | 96 (43%) | 0.010** |
| *Household owns watch* | 89 (44%) | 130 (59%) | 0.002*** |
| **Other respondent characteristics** |  |  |  |
| Respondent completed primary school or above | 128 (63%) | 140 (63%) | 0.949 |
| Respondent completed secondary school or above | 18 (9%) | 33 (15%) | 0.060* |
| Respondent has moderate problems walking about, or worse | 12 (6%) | 13 (6%) | 0.971 |
| Respondent has moderate pain or discomfort, or worse | 21 (10%) | 17 (8%) | 0.325 |
| Respondent rents dwelling | 60 (30%) | 54 (24%) | 0.213 |
| Respondent's dwelling has zinc or concrete roof | 202 (100%) | 222 (100%) | n/a |
| **Compound-level WASH characteristics** |  |  |  |
| Water available at least 8 hours / day | 99 (49%) | 110 (50%) | 0.912 |
| Uses on-plot toilet | 197 (98%) | 219 (99%) | 0.397 |
| Shares toilet with other household(s) | 181 (90%) | 196 (88%) | 0.667 |
| Number of households sharing stance | 3.3 (1.7) | 3.2 (1.6) | 0.511 |
| Number of people sharing stance | 11.8 (5.2) | 12.6 (6.6) | 0.170 |

Data are n (%) for categorical variables and mean (SD) for numerical variables. \*, \*\*, \*\*\* indicate significance at the 10, 5 and 1 percent level. Variables included in the wealth index are italicised. One participant had missing data for the wealth index. In the replication dataset, we categorised age, household size and children under 14 to maintain full anonymity, since several values were shared by 5 people or fewer. This table reports the mean of continuous values.

For hypotheses 1b and 1c, we estimated a GLMM with the same covariates but a different dependent variable as appropriate. Likewise for hypothesis 3 we regressed on SanQoL attribute-level scores, which range from 0 to 3 (Online Appendix B). For hypotheses 2a and 2b, we specified the same models but including a factorial interaction with *T* for the gender or elderly binary variable.

We assessed the sensitivity of results as follows. First, we included in ***X***_*ij*_ only covariates significantly different between groups at the 10% level (Table 2) and excluded the gender and aged 60+ binary variables. Second, to explore the presence of omitted variable bias, we included additional covariates with the potential to influence SanQoL (gender, aged 60+, renting dwelling, number of people sharing a toilet stance, whether the toilet is shared with other households). Third, to explore omitted variable bias in the WHO-5 regressions, we included covariates hypothesised to affect mental wellbeing (gender, aged 60+, having a partner, being in moderate pain, or having moderate problems walking). Fourth, we explored whether using a generalised estimating equations (GEE) or ordinary least squares (OLS) specification instead of GLMM affected the results. Finally, we explored whether using mixed-effects ordered logit for the SanQoL attribute-level regressions affected results, instead of treating them as continuous variables. We include the STROBE checklist in Online Appendix G,^30^ and a reflexivity statement with checklist in Online AppendixH.^31^

### Ethical approval

The study received prior approval from the *Comité Nacional de Bioética para a Saúde* (IRB00002657) at the Ministry of Health in Mozambique, and the Research Ethics Committee of the London School of Hygiene and Tropical Medicine (Ref: 14609). Informed, written consent was obtained from all participants.

### Patient and public involvement

Members of the public were not involved in the design or conduct of this specific study. However, members of the public were involved in development of the SanQoL outcome measure as: (i) participants in the qualitative research informing attribute identification;^10^ (ii) participants in the piloting and cognitive interviews informing item development.^20^

## 3. Results

We sampled individuals from 424 different households across 272 compounds (clusters), of which 130 were control and 142 intervention, during April-May 2019 (Figure 1). In some compounds, only one man or woman was eligible (mean respondents per cluster: 1.6), which did not affect our study objectives. The response rate amongst eligible participants was 99%. There was no evidence of difference in background characteristics between intervention/control at the 5% level, except for the wealth index score (Table 2). People living in intervention compounds were slightly wealthier than controls, but assets that were different were the less expensive ones (e.g. watch, radio).

The intervention was associated with an adjusted gain in SanQoL of 0.34 (95% CI: 0.29-0.38), noting that SanQoL is measured on a 0-1 scale and the control mean was 0.49 (Table 3). Full regression results are in Online Appendix D. The effect size was very large at 1.6 standard deviations (SD). Comparing effects between the two intervention toilet designs, there was weak evidence (p=0.079) for users of STs having higher SanQoL than users of CSBs (Online Appendix D, Table D-2). Measured on the sanitation VAS, which is scored 0-10, the intervention was associated with a 2.9 point gain (95% CI: 2.4 - 3.4)(Table 3). The effect size was very large at 1.3 SD, similar to that of SanQoL. Considering WHO-5 mental wellbeing, measured on a 0-100 scale, the intervention was associated with a 6.2 point gain (95% CI: 0.3-12.2)(Table 3). The effect size was small at 0.2 SD.

**Table 3:** Effects on primary and secondary outcomes

| Outcome | Means |  | Unadjusted models |  | Adjusted models |  |  |
| --- | --- | --- | --- | --- | --- | --- | --- |
|  | Control<br>(n=202)<br>Mean (SE) | Interv'n<br>(n=222)<br>Mean (SE) | Unadjusted<br>difference<br>(95% CI) | p-value | Adjusted<br>difference<br>(95% CI) | p-value | Adjusted<br>effect size<br>(Cohen's d) |
| SanQoL<br>(0-1 scale) | 0.49<br>(0.02) | 0.83<br>(0.01) | 0.34***<br>(0.29 - 0.38) | <0.001 | 0.34***<br>(0.29 - 0.39) | <0.001 | 1.6 |
| Sanitation<br>VAS<br>(0-10 scale) | 4.1<br>(0.2) | 7.0<br>(0.1) | 2.9***<br>(2.4 - 3.4) | <0.001 | 2.9***<br>(2.4 - 3.4) | <0.001 | 1.3 |
| WHO-5<br>(0-100 scale) | 54.4<br>(1.9) | 58.7<br>(1.9) | 5.6*<br>(-0.4 - 11.6) | 0.065 | 6.2**<br>(0.3 - 12.2) | 0.041 | 0.2 |

Adjusted models include gender, aged over-60, and wealth score as covariates. Standard errors are clustered at the compound level. \*, \*\*, \*\*\* indicate significance at the 10, 5 and 1 percent level. Detailed regression output is in Online Appendix D.

With the caveat that our study was not powered for sub-group analyses, interaction terms provided no evidence for any outcome that women benefitted more from better toilets than men (Online Appendix D, Table D-1). Regardless of the intervention, there was also no evidence that women reported lower SanQoL than men in the sample as a whole, though there was weak evidence (p=0.056) that this was the case for the VAS. Considering elderly people as a sub-group, there no evidence that people aged over 60 benefitted more from better toilets than under-60s (Online Appendix D, Table D-1).

In regressions on individual attribute scores, the intervention was associated with gains across all five SanQoL attributes (Table 4). Effect sizes were largest for safety (1.5 SD) and privacy (1.4 SD). Coefficients on interaction terms for gender and aged over 60 were not significant, as with SanQoL index values.

**Table 4:** Effects on individual SanQoL attribute scores (ranging 0-3) and interactions with gender and age

|  |  | Disgust | Health | Shame | Safety | Privacy |
| --- | --- | --- | --- | --- | --- | --- |
| Means | Control (n=202)<br>Mean (SE) | 1.59<br>(0.082) | 1.40<br>(0.085) | 1.56<br>(0.081) | 1.29<br>(0.080) | 1.58<br>(0.081) |
|  | Intervention (n=222)<br>Mean (SE) | 2.32<br>(0.067) | 2.36<br>(0.060) | 2.40<br>(0.068) | 2.64<br>(0.044) | 2.84<br>(0.037) |
| Main model<br>(unadjusted) | Unadjusted<br>difference (95% CI) | 0.72<br>(0.50 - 0.94) | 0.96<br>(0.74 - 1.18) | 0.82<br>(0.6 - 1.04) | 1.35<br>(1.16 - 1.54) | 1.26<br>(1.07 - 1.45) |
|  | p-value | <0.001*** | <0.001*** | <0.001*** | <0.001*** | <0.001*** |
| Main model<br>(adjusted) | Adjusted difference<br>(95% CI) | 0.75<br>(0.53 - 0.97) | 0.96<br>(0.74 - 1.18) | 0.80<br>(0.58 - 1.02) | 1.36<br>(1.16 - 1.56) | 1.25<br>(1.06 - 1.44) |
|  | p-value | <0.001*** | <0.001*** | <0.001*** | <0.001*** | <0.001*** |
|  | Adjusted effect size<br>(Cohen's d) | 0.7 | 0.9 | 0.7 | 1.5 | 1.4 |
| Gender<br>interaction<br>model<br>(adjusted) | p-value on<br>coefficient for<br>Female*Intervention | 0.56 | 0.98 | 0.19 | 0.29 | 0.83 |
| Over-60<br>interaction<br>model<br>(adjusted) | p-value on<br>coefficient for Over-60*Intervention | 0.43 | 0.28 | 0.87 | 0.15 | 0.54 |

Models include gender, aged over-60, and wealth score as covariates. Standard errors are clustered at the compound level. \*, \*\*, \*\*\* indicate significance at the 10, 5 and 1 percent level. Detailed regression output is in Online Appendix D.

In the first robustness check, including only covariates significantly different between groups at the 10% level (Table 2), there was no meaningful difference to results for any of the three outcomes (Online Appendix E, Tables E-1 and E-2). Second, when all covariates hypothesised as influencing SanQoL and VAS were included, there was no evidence of omitted variable bias in terms of the size and p-value of the coefficient on the intervention variable. However, the coefficient on the binary covariate for sharing the toilet was negative and significant at the 1% level in both SanQoL and VAS regressions. This finding indicates that sharing a toilet with other households makes people worse off, and is explored as a factorial interaction in Online Appendix F. Third, when all covariates hypothesised as influencing mental wellbeing were included in the WHO-5 regression, there was no evidence of omitted variable bias (Table E-2). Fourth, using a GEE or OLS specification did not affect headline results for SanQoL or VAS (Table E-1). The effect on WHO-5 was significant only at the 10% level in the OLS regression (Table E-2), but OLS is unlikely to be appropriate for the hierarchical structure of our data. Furthermore, residuals were bimodally distributed in the WHO-5 OLS regression, suggesting the GLMM and GEE specifications are preferred. Finally, using mixed-effects ordered logit for SanQoL attribute-level regressions, rather than treating them as continuous variables in GLMM, made no difference to interpretation (Table E-3). We conclude from our robustness checks that our main models were appropriate for testing our hypotheses.

## 4. Discussion

In this observational study building on the design of the earlier MapSan trial, we find that users of high-quality shared toilets experienced a 1.6 SD gain in sanitation-related quality of life compared to pit latrines, and a 0.2 SD gain in broader mental wellbeing. This intervention had no effect on under-5 health outcomes,^27^ so our findings demonstrate that better toilets can improve people’s lives beyond infectious disease, at a time when several randomised trials have questioned the health effects of sanitation improvements.^21^

Since all people in intervention compounds were previously sharing a low-quality toilet with the same people in the same location, the mechanism driving our results is likely to be the specific characteristics of intervention toilets. Solid walls and doors likely improved perceptions of privacy, safety and shame compared to PLs with makeshift walls and doors (photos in Online Appendix A). The pour-flush interface was likely to have reduced smells and visible faeces compared to PLs without water seals, improving perceptions of disgust, shame, and health risk. Users value such toilet characteristics – a choice experiment in urban Zambia found willingness to pay (WTP) additional rent for solid toilet doors was 8% of median monthly rent, and WTP for flush toilets as opposed to pit latrines was 5% of rent.^32^

While it is intuitive that people using better-quality toilets experience more privacy or less disgust, our contribution is in quantifying this to inform decisions based on comparative effectiveness, which has not previously been done.^33^ The fact that SanQoL is specific to sanitation limits its broader relevance. However, such “condition-specific” outcomes focused on experienced symptoms (e.g. of arthritis or asthma) within only a few QoL domains are regularly used to evaluate interventions targeting those specific problems.^34^ The small effect on mental wellbeing was expected, as it is a more distal outcome than SanQoL. A previous cross-sectional study identified associations of urban sanitation access with WHO-5,^22^ and our contribution is in evaluating a specific urban sanitation intervention.

Despite willingness to contribute financially to 10-15% of capital costs being an enrolment criterion for both intervention and control in MapSan, it is possible that wealthier people were more likely to uptake the intervention due to being able to afford this contribution. Since our survey was four years after the intervention, any differences could be as a result of the intervention. However, any wealth effect would likely be in the other direction since intervention households reported spending substantially more than controls on both cleaning and maintenance.^35^

Limitations of our study include that we relied on the controlled before-and-after design of a previous trial in which the intervention was not randomly allocated, risking selection bias. Our design necessitates adjusting for covariates which may be imprecisely measured, and the absence of pre-intervention SanQoL data precluded adjustment for baseline values. While our comparison groups were well-balanced overall, and we adjusted for the only unbalanced covariate, we cannot account for unobserved confounding. We excluded people who had lived in the compound for less than four years to account for in-migration since the intervention. However, risk of bias from out-migration remains, for example if people with certain characteristics were more likely to have moved away. Overall, the magnitude of the effect size for our primary outcome (1.6 SD) means it is unlikely to be explained by bias alone.

As with any subjective self-reported outcome, there is risk of reporting bias which is difficult to account for, though we assume that any measurement error was not correlated with toilet type. In introducing themselves, fieldworkers emphasised that they were not linked to the implementing NGO or government, but intervention respondents may have wanted to appear grateful and control respondents may have wanted to appear badly off. Reporting bias could pose more of a risk to the mental wellbeing finding with its higher p-value, but the WHO-5 questions do not refer to sanitation in any way, so may have been perceived by respondents as being less linked to the intervention.

Since physical environments and sanitary conditions in these parts of Maputo are similar to large portions of other Mozambican cities, it is likely that findings would be generalisable to those settings, as well as parts of other cities in many African countries. However, social environments differ within and beyond Mozambique, and are likely to influence the relationships explored. Future intervention trials should include QoL outcomes such as SanQoL, since these benefits are highly valued by users. Changes in sanitation-specific QoL outcomes such as privacy and disgust are likely to suffer from fewer confounding factors than infectious disease outcomes, since they are more proximal to the service being provided.

## 5. Conclusion

Quality of life outcomes highly valued by toilet users and can be improved by sanitation interventions. If applied in future impact evaluations, SanQoL, WHO-5 and similar measures could help sanitation decision-makers understand which types of sanitation interventions most improve people’s lives. QoL indices with weighting derived from respondent valuation tasks, such as SanQoL, can also be used in economic evaluation to identify interventions which are most efficient use of resources, not only those which are most effective. The likelihood of positive health externalities remains a core rationale for public investment in sanitation. However, in the almost certain absence of intervention-specific data on health effects, policy-makers are likely to be willing to make more informed decisions on the basis of QoL outcomes which are more easily and quickly measured in their specific setting.

## Data Availability

Deidentified individual participant data, data dictionary and replication code are available open access on the LSHTM data repository.

https://doi.org/10.17037/DATA.00002442

## Acknowledgements

We greatly appreciated the cooperation of survey respondents. We also appreciated the efforts of the fieldworkers employed by WE Consult: Euclimia Titosse, Carla Panguene, João-Pedro Guambe and Faustino Benzane. The authors gratefully acknowledge the technical and logistical support received from Vasco Parente and his team at WSUP Mozambique. We benefitted from the comments of Roxanne Kovacs and seminar participants at the Global Health Economics Centre at LSHTM, as well as from Britta Augsburg and Antonella Bancalari.

## Data availability statement

Deidentified individual participant data, data dictionary and replication code are available open access on the LSHTM data repository: https://doi.org/10.17037/DATA.00002442.

