## Supplementary material for "Impact of a sanitation intervention on quality of life and mental wellbeing in low-income urban neighbourhoods of Maputo, Mozambique": Online Appendices

A. Additional information on setting and intervention
B. Additional methods
C. Distributions of outcome variables
D. Additional results
E. Robustness checks
F. The role of sharing toilets
G. STROBE checklist
H. Reflexivity statement

### A. Additional information on setting and intervention

#### *Photographs with typical examples of main toilet types*

Below are photographs of typical toilets of each type. CSB and ST designs are fairly homogenous, with some variation in the type of squat plate or seat pan used. The level of sanitation service they provide is the same, though CSBs also have a rooftop water tank and two laundry stations. Pit latrines are far more diverse. Some may nominally meet the WHO/UNICEF Joint Monitoring Programme's definition of an improved technology (e.g. photo 2 has a concrete slab). Such latrines may therefore be categorised as "limited" sanitation (since they are shared) rather than "unimproved".

*Figure A-1: Pit latrines (control)*

| 1. Pit latrine with tyre and wood for squatting | 2. Pit latrine with concrete slab |
|---|---|
| 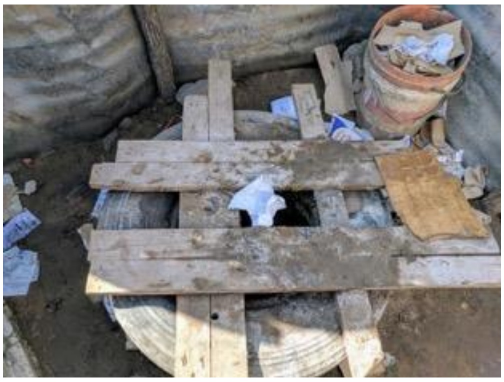 | 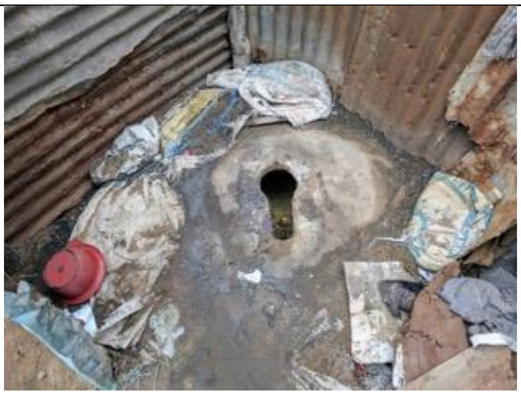 |
| 3. Fabric door providing limited privacy | 4. No door and adjacent greywater pit |
| 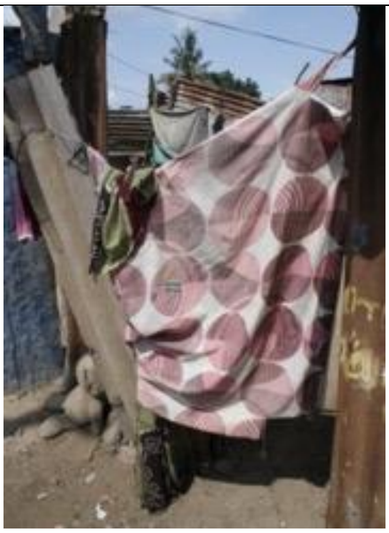 | 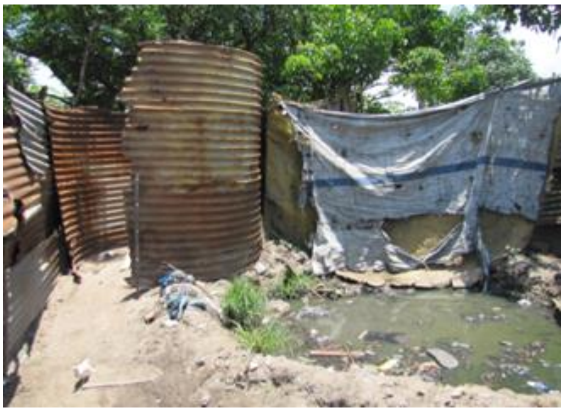 |

*Figure A-2: Shared toilets and community sanitation blocks (intervention)*

<table>
<tr><th colspan="2">Exterior</th></tr>
<tr><td>1. Shared toilet (ST)</td><td>2. Community sanitation block (CSB)</td></tr>
<tr><td>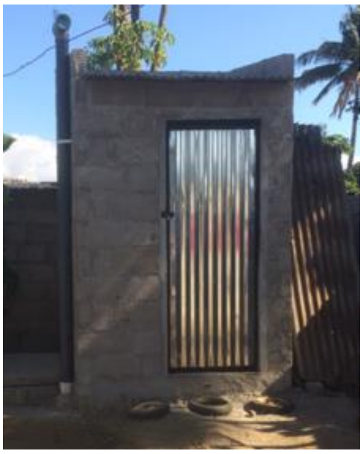</td><td>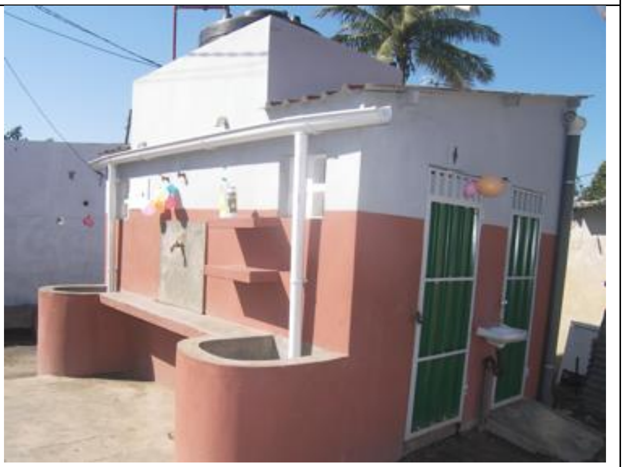</td></tr>
<tr><th colspan="2">Interior (varied between CSB / ST depending on design)</th></tr>
<tr><td>3. Squat pan</td><td>4. Seat pan</td></tr>
<tr><td>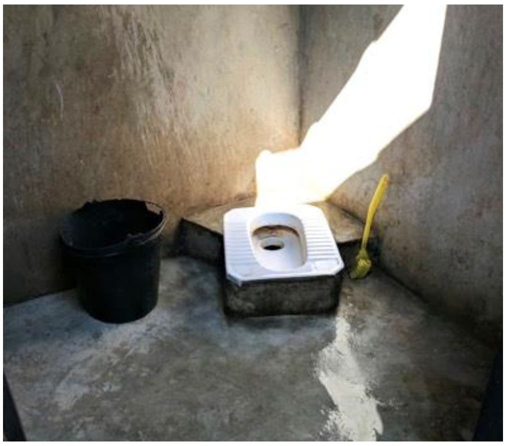</td><td>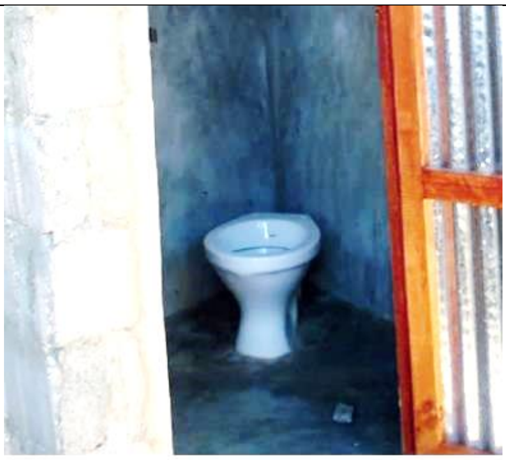</td></tr>
</table>

*Map of respondent households within Maputo*

Figure A-3 Panel A shows the greater Maputo region which, including the adjoining city of Matola, has a population of 2.9 million.[1] Panel B shows the geolocations of households included in our survey (n=424). They are situated within a small area of about 10km$^2$ within the Nhlamankulu district. Since compounds were randomly sampled from the list of MapSan-enrolled compounds, this broadly represents the implementation area of the intervention overall.

*Figure A-3: Maps of Maputo*

A. Greater Maputo region

B. Respondent households within Maputo City

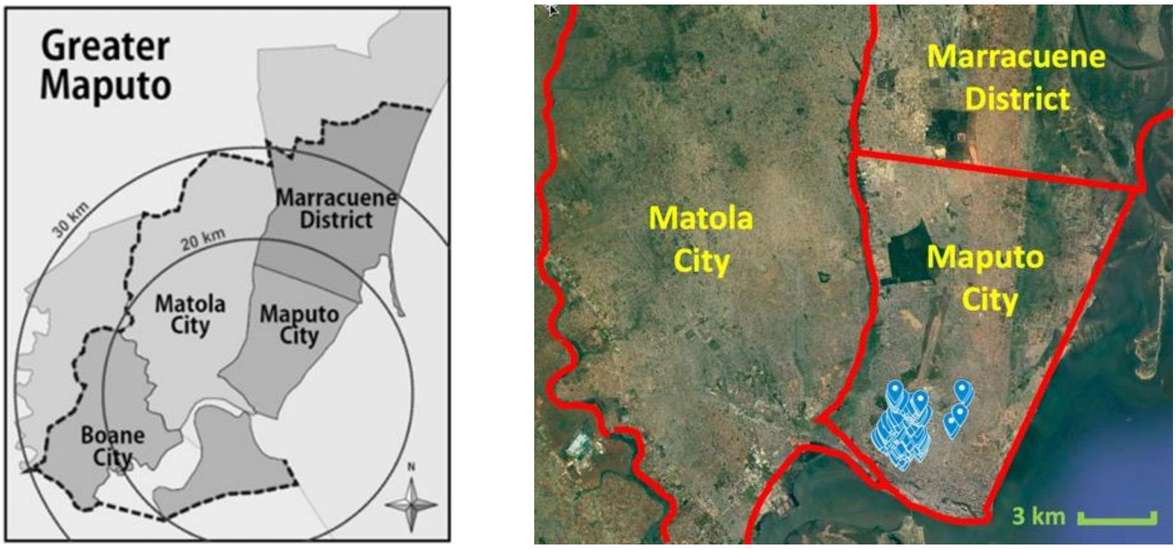


Source: Batran et al. (2018)[2]

Source: Google Earth

*Additional information on intervention delivery*

The roles of key stakeholders involved in intervention delivery are summarised below. Further information is provided elsewhere.[3,4]

*Table A-1: Stakeholders involved in intervention delivery*

| Stakeholder | Overall role | Key activities |
|---|---|---|
| WSUP (NGO) | Provider and project lead | • Project design and management<br>• Manage design consultants<br>• Manage construction contractors<br>• Supervise construction |
| Community-based organisations | Sub-contractor facilitating community engagement | • Facilitate site selection<br>• Collect household capital contribution |
| Construction firms | Sub-contractors constructing the toilet infrastructure | • Dismantle old toilet<br>• Construct new toilet |
| Households | User of infrastructure | • Contribute 10-15% of capital costs<br>• Clear site of material<br>• Participate in meetings and data collection |
| Municipality (*Conselho Municipal de Maputo*, CMM) | Oversight and approvals by department for water and sanitation | • Approve designs and procurement<br>• Provide permits for CSBs<br>• Monitor infrastructure |
| World Bank | Oversight of overall programme | • Fund overall project<br>• Oversight of delivery |

*Table A-2: Intervention description using TIDieR checklist (Hoffman et al. 2014)*[5]

| | **Item** | **Notes** |
|---|---|---|
| **1** | Provide the name or a phrase that describes the intervention. | Subsidised pour-flush toilets shared by multiple households |
| **2** | Describe any rationale, theory, or goal of the elements essential to the intervention. | In this setting there is limited space and willingness or ability to pay for private toilets, and households already use low-quality shared pit latrines. |
| **3** | Materials: Describe any physical or informational materials used in the intervention, including those provided to participants or used in intervention delivery or in training of intervention providers. Provide information on where the materials can be accessed (e.g. online appendix, URL). | The intervention provided two types of toilet facility (photos above), alongside education on their use and maintenance. There were also two hygiene promotion visits after completion of construction, carried out by paid staff who received 2 days of training. These focused on contamination routes, good personal hygiene practice, and handwashing with soap. More information is provided elsewhere.[3,4,6] |
| **4** | Procedures: Describe each of the procedures, activities, and/or processes used in the intervention, including any enabling or support activities. | Key procedures included:<br>1. Community engagement and site identification – undertaken by eight contracted community-based organisations (CBOs), e.g. assessment of |

| | | |
|---|---|---|
| | | demand for better toilets and localised environmental issues affecting site selection (e.g. water table)<br>2. Site selection and preparation – site selection undertaken by WSUP in discussion with CBOs, and site preparation (e.g. emptying of old latrine pits) undertaken by contracted firms.<br>3. Toilet construction – undertaken by contracted construction firms<br>4. Education on use, maintenance and hygiene – undertaken by contracted 'sanitation activists' |
| **5** | For each category of intervention provider (e.g. psychologist, nursing assistant), describe their expertise, background and any specific training given. | Main stakeholders in delivery included:<br>1. **Water and Sanitation for the Urban Poor** (international NGO) – overall lead on intervention delivery. Team included engineers and community engagement specialists.<br>2. **Various community-based organisations** – sub-contractor facilitating community engagement. 48 people trained. Teams included facilitators from the local area of the intervention.<br>3. **Various construction firms** – Sub-contractors building the toilet infrastructure. They were predominantly small local firms.<br>4. **Sanitation activists** – Sub-contractors educating toilet users and promoting hygiene. 55 people trained.<br>5. **Municipality and World Bank** – oversight and approvals. Team included engineers. |
| **6** | Describe the modes of delivery (e.g. face-to-face or by some other mechanism, such as internet or telephone) of the intervention and whether it was provided individually or in a group. | All engagement was face-to-face. As this was shared sanitation, any site visits were made to compound members jointly, rather than individually. |
| **7** | Describe the type(s) of location(s) where the intervention occurred, including any necessary infrastructure or relevant features. | Setting described fully in manuscript main body. |
| **8** | Describe the number of times the intervention was delivered and over what period of time including the number of sessions, their schedule, and their duration, intensity or dose. | All aspects of the intervention delivered only once, except for two hygiene promotion visits. |
| **9** | If the intervention was planned to be personalised, titrated or adapted, then describe what, why, when, and how. | n/a |
| **10** | If the intervention was modified during the course of the study, describe the changes (what, why, when, and how). | n/a |
| **11** | Planned: If intervention adherence or fidelity was assessed, describe how and by whom, and if any strategies were used to maintain or improve fidelity, describe them. | n/a |
| **12** | Actual: If intervention adherence or fidelity was assessed, describe the extent to which the intervention was delivered as planned. | Fidelity was assessed by Bick et al.[4] |

### B. Additional methods

#### *Sampling of individuals*

Upon arrival at the next compound on the list, and with the approval of a resident, fieldworkers inspected the toilet to assess eligibility. Next, by talking to residents, they listed all eligible people based on the inclusion criteria. For the male respondent, sampling was random from the list of eligible men within the compound, by approaching households starting from the house opposite the compound entrance, and working leftwards until an eligible man was identified. The same process was then followed for eligible women, with the condition that the female respondent not be from the same household as the male respondent. We continued visiting compounds until the target sample size was reached. Interviews were in Portuguese, unless the respondent preferred to talk in Changana, a local language in which all interviewers were fluent. We collected data on smartphones using the mWater surveyor application.

*Table B-1: Hypotheses*

| | Hypothesis | Rationale |
|---|---|---|
| **1. Main** | (a) the intervention is associated with higher SanQoL index values | The intervention should improve people's sanitation-related capabilities and self-perceived level of sanitation in general.[7,8] |
| | (b) the intervention is associated with higher sanitation VAS score | |
| | (c) the intervention is associated with higher mental wellbeing (WHO-5) | Sanitation interventions may improve people's self-perceived mental wellbeing, as shown in a systematic review of mainly qualitative studies,[9] and cross-sectional studies using WHO-5.[10,11] |
| **2. Sub-groups** | (a) across all three outcomes, any intervention effect is larger for women than men | Women experience sanitation differently to men in that they squat for urination, undertake menstrual hygiene, and are more likely to fear and experience assault.[12] Sanitation deficits particularly impact on women,[13] so improvements in sanitation may disproportionately benefit women more. |
| | (b) across all three outcomes, any intervention effect is larger for elderly people (aged 60+) than non-elderly | Elderly people are impacted by poor sanitation, predominantly as a result of disabilities that occur with aging.[14] Therefore, improvements in sanitation may disproportionately benefit elderly people. In Mozambique, an elderly person is defined in law as anyone aged 60 years or older.[15] |

#### *Ranking exercise*

The ranking exercise comprised a velcro-covered A4 plastic board with a 30cm vertical line and 10 intervals from 1-10 marked, as well as velcro-backed laminated cards (Figure B-1). The exercise was based on methods reported in Drummond et al.[16] Each of the cards was labelled with attributes emerging from prior qualitative work, which were already familiar to respondents from the earlier parts of the questionnaire. Participants were asked to place the cards on the scale according to their relative importance. They were first asked to choose the card representing the attribute they thought most important for a good toilet and a good life. They were asked not to focus on their current toilet but consider their ideal toilet. They were then asked to do the same for the least important attribute of the remaining cards. These were placed at the top (10) and bottom (1) of the board. The enumerator explained that being at position 1 does not mean that attribute is not important, but just that it is *less* important than the others. The respondent was then asked to stick the remaining cards to the board, at the places on the line that they consider reflected relative importance. Moving attributes was allowed. Placing more than one attribute at the same position was also allowed (only 5 out of 424 participants did so), which is accounted for in Equation A such that weights still sum to 1.

*Figure B-1: SanQoL attributes ranking board*

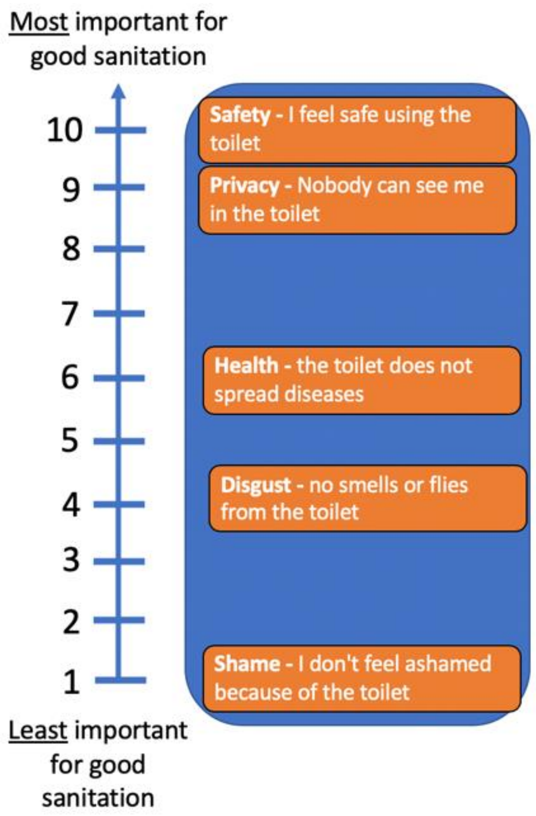


Using data on mean ranks, we estimated weights (Equation A) for each SanQoL dimension using the rank sum method,[17] as previously used in index valuation.[18,19] Mean ranks are reported elsewhere,[20] as are differences in attribute ranks by gender, aged 60+, and treatment.

*Equation A – attribute weights for a population*

$$w_i = \frac{N - R_i + 1}{\sum_{i=1}^{N}(N - R_i + 1)}$$

*Equation B – SanQoL index value for an individual*

$$S_j = \frac{\sum_{i=1}^{N}(x_{ij} * \mathrm{w}_i)}{3}$$

where:

- $w_i$ is the weight of the ith attribute
- $N$ is the number of attributes
- $R_i$ is the mean rank of the ith attribute in the population
- $x_{ij}$ are item scores ranging from 0-3 for the jth individual, where "always"=3 and "never"=0.
- $S_j$ is the SanQoL index value for the jth individual

#### *Visual analogue scale (VAS)*

The sanitation VAS was adapted from the VAS in the EQ-5D measure of health-related quality of life,[21] with emoji visualisation informed by the visual pain scale.[22] The enumerator reads out the guidance (Figure B-2) then the respondent indicates their selected level on the scale with a pencil. The VAS gives us information about people's overall assessment of their level of sanitation, while SanQoL restricts the evaluative space to five attributes with population-based weights. The VAS therefore measures something conceptually different, but complementary, to SanQoL.

*Figure B-2: sanitation VAS*

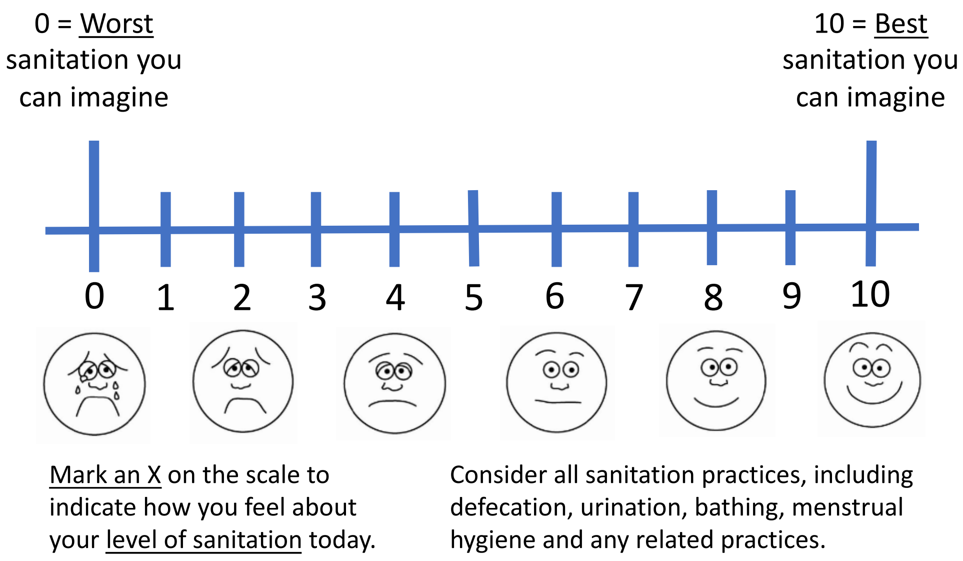

##### *WHO-5 attributes*

WHO-5 attributes are listed in the table below. The raw score ranges from 0 to 25, and is multiplied by 4 to reach a score where 100 represents best possible mental wellbeing and 0 worst possible.

*Table B-2: WHO-5 attributes*

| # | Attribute | Item | Responses |
|---|---|---|---|
| 1 | Cheerful | In the last 2 weeks have you felt cheerful and in good spirits? | 0 - At no time<br>1 - Some of the time<br>2 - Less than half of the time<br>3 - More than half of the time<br>4 - Most of the time<br>5 - All of the time |
| 2 | Calm | In the last 2 weeks have you felt calm and relaxed? | |
| 3 | Active | In the last 2 weeks have you felt active and vigorous? | |
| 4 | Fresh | In the last 2 weeks have you woken up feeling fresh and rested? | |
| 5 | Interest | In the last 2 weeks have you had a daily life filled with things that interest you | |

### C. Distributions of outcome variables

*Figure C-1: Histograms of primary and secondary outcomes by toilet type*

Panel 1 - SanQoL index values

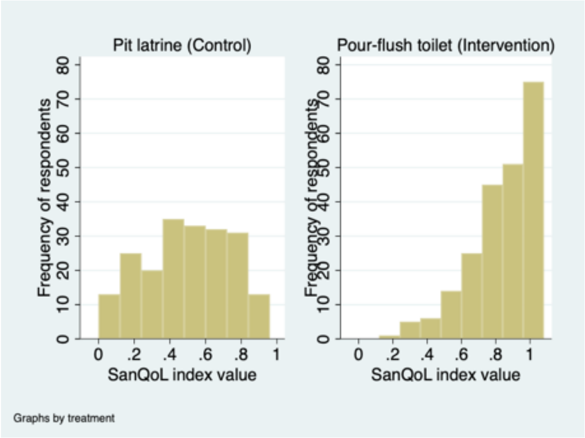


Panel 2 – Sanitation VAS

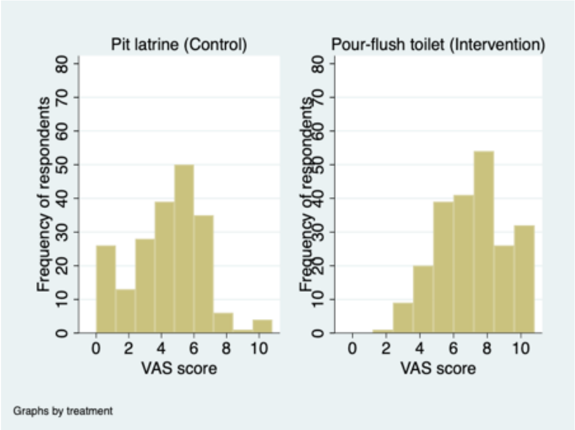


Panel 3 – WHO-5 index

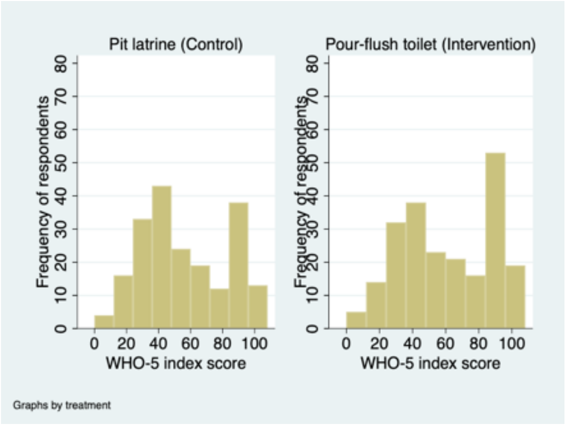


*Figure C-2: Distributions of SanQoL attributes by intervention and control*

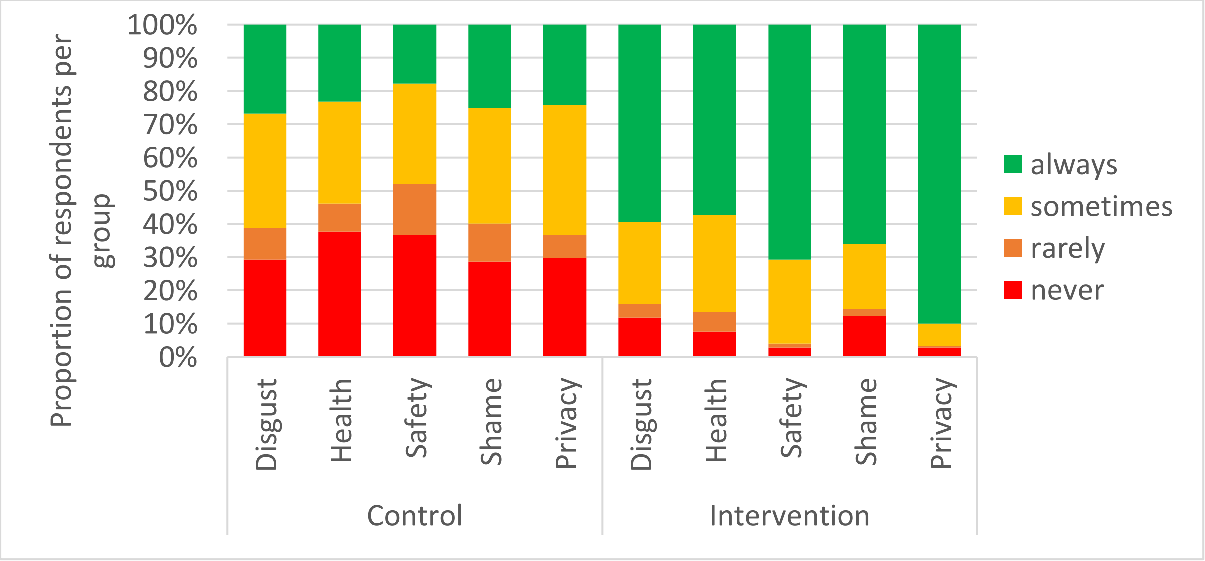


Note: Scores range from 0-3 representing a scale from never to always (Table 1).

*Table C-1: Polychoric inter-item correlations for SanQoL attributes*

| | Disgust | Health | Shame | Safety | Privacy |
|---|---|---|---|---|---|
| **Disgust** | 1.00 | | | | |
| **Health** | 0.56 | 1.00 | | | |
| **Shame** | 0.52 | 0.53 | 1.00 | | |
| **Safety** | 0.41 | 0.47 | 0.49 | 1.00 | |
| **Privacy** | 0.40 | 0.43 | 0.54 | 0.70 | 1.00 |

*Table C-2: SanQoL questions in Portuguese*

| # | Dimension | Question | Responses |
|---|---|---|---|
| 1 | Disgust<br>*Nojo* | Can you use the toilet without feeling disgusted?<br>*Pode usar a casa de banho sem se sentir nojo?* | 3 – Always (*sempre*)<br>2 – Sometimes (*as vezes*)<br>1 – Rarely (*raramente*)<br>0 – Never (*nunca*) |
| 2 | Health<br>*Saúde* | Can you use the toilet without worrying that it spreads diseases?<br>*Pode usar a casa de banho sem se preocupar que espalhe doenças?* | |
| 3 | Privacy<br>*Privacidade* | Can you use the toilet in private, without being seen?<br>*Pode usar a casa de banho com privacidade, sem ser visto/a?* | |
| 4 | Shame<br>*Vergonha* | Can you use the toilet without feeling ashamed for any reason?<br>*Pode usar a casa de banho sem sentir vergonha por qualquer motivo?* | |
| 5 | Safety<br>*Segurança* | Are you able to feel safe while using the toilet?<br>*É capaz de se sentir seguro/a ao usar esta casa de banho?* | |

### D. Additional results

*Table D-1: Regression output underlying Tables 4 and 5*

| | SanQoL index value | | | Sanitation-VAS | | | WHO-5 index | | |
|---|---|---|---|---|---|---|---|---|---|
| | (1) main regression | (2) gender interaction | (3) age interaction | (4) main regression | (5) gender interaction | (6) age interaction | (7) main regression | (8) gender interaction | (9) age interaction |
| Intervention toilet | 0.34***<br>(0.02) | 0.33***<br>(0.03) | 0.34***<br>(0.02) | 2.91***<br>(0.24) | 2.76***<br>(0.30) | 2.91***<br>(0.26) | 6.25**<br>(3.05) | 6.65*<br>(3.80) | 6.78**<br>(3.22) |
| Aged 60+ | -0.01<br>(0.03) | -0.01<br>(0.03) | -0.03<br>(0.05) | -0.15<br>(0.27) | -0.12<br>(0.28) | -0.14<br>(0.45) | -12.95***<br>(2.91) | -13.03***<br>(2.95) | -10.64**<br>(4.20) |
| Female | -0.01<br>(0.02) | -0.02<br>(0.03) | -0.01<br>(0.02) | -0.30*<br>(0.16) | -0.45*<br>(0.24) | -0.30*<br>(0.16) | -3.31*<br>(1.94) | -2.91<br>(2.56) | -3.47*<br>(1.98) |
| Wealth index score | -0.01<br>(0.01) | -0.01<br>(0.01) | -0.01<br>(0.01) | -0.07<br>(0.10) | -0.07<br>(0.10) | -0.07<br>(0.10) | 0.98<br>(1.12) | 0.98<br>(1.12) | 1.00<br>(1.11) |
| Intervention toilet * female | | 0.03<br>(0.04) | | | 0.29<br>(0.33) | | | -0.77<br>(3.88) | |
| Intervention toilet * aged 60+ | | | 0.03<br>(0.06) | | | -0.02<br>(0.57) | | | -4.09<br>(5.69) |
| Constant | 0.50***<br>(0.02) | 0.51***<br>(0.03) | 0.50***<br>(0.02) | 4.28***<br>(0.20) | 4.36***<br>(0.22) | 4.28***<br>(0.20) | 57.01***<br>(2.40) | 56.82***<br>(2.53) | 56.83***<br>(2.40) |
| Observations | 423 | 423 | 423 | 423 | 423 | 423 | 422 | 422 | 422 |

Note: Cells report regression coefficients, with standard errors (clustered at compound level) in parentheses. *, **, *** indicate significance at the 10, 5 and 1 percent level. SanQoL is on a 0-1 scale, VAS is on a 0-10 scale, and WHO-5 is on a 0-100 scale.

Residuals are approximately normally distributed for all three main results (Figure D-1), and plots of residuals against fitted values for the fixed portions raise no concerns about heteroscedasticity. For the SanQoL plot (panel 4) the plot for the intervention group appears truncated, due to the modal SanQoL index value being 1, which effectively censors the residuals.

*Figure D-1: Histograms of residuals and residuals-vs-fitted plots for main results in Table 3 (column references are to Table D-1)*

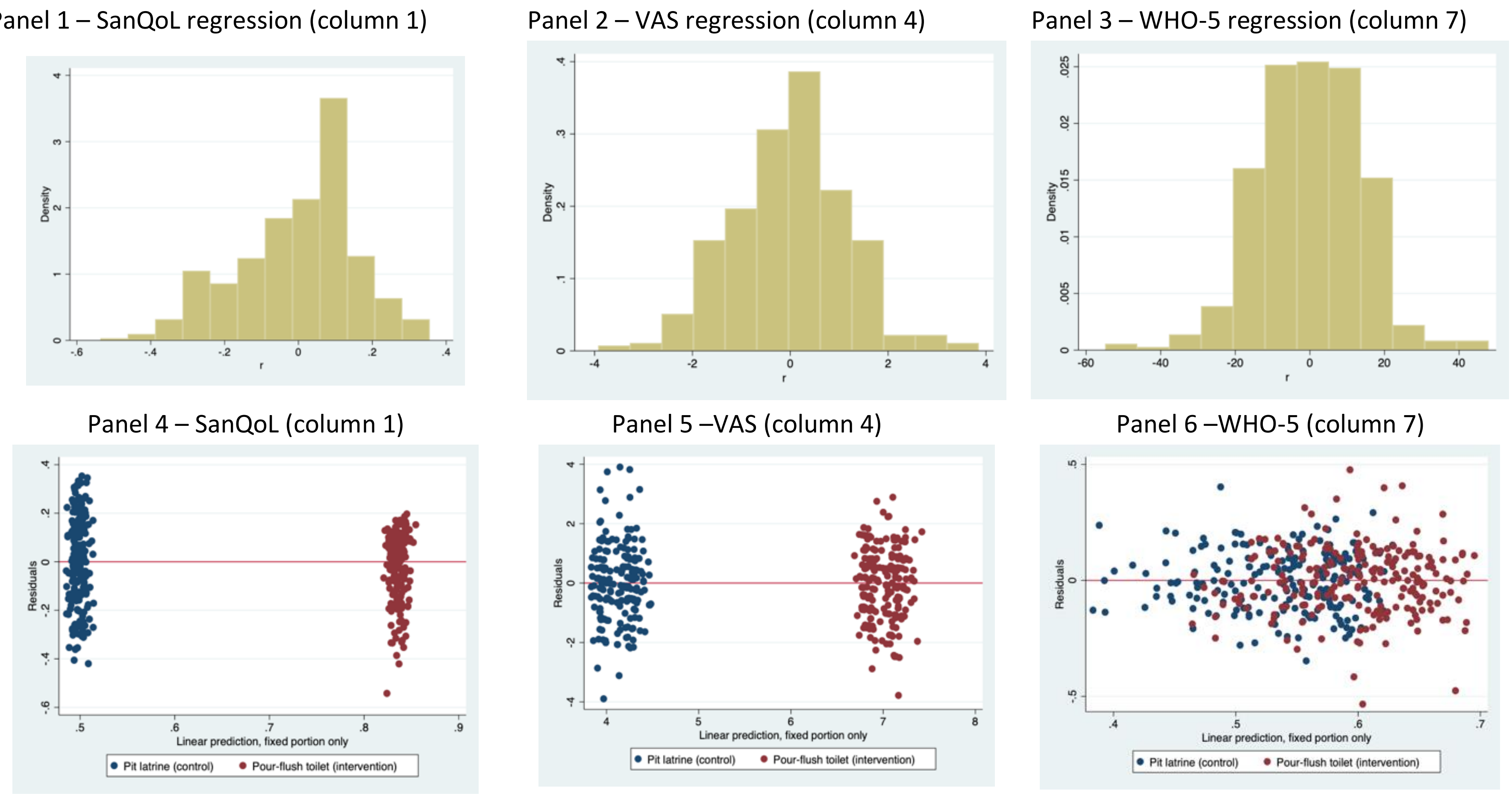

Below is the regression output for the main results regressions on SanQoL index values, but including toilet type as a covariate to estimate whether there was a difference between shared toilets (ST) and community sanitation blocks (CSB). It shows that there is weak evidence (p=0.079) for a larger effect of the intervention on ST users than CSB users, which is important for subsequent cost-effectiveness analysis.

*Table D-2: Regression output including toilet type as covariate*

| | |
|---|---|
| Intervention toilet | 0.28***<br>(0.04) |
| Aged 60+ | -0.02<br>(0.03) |
| Female | -0.01<br>(0.02) |
| Wealth index score | -0.01<br>(0.01) |
| Toilet type = ST | 0.068*<br>(0.04) |
| Constant | 0.50***<br>(0.02) |
| Observations | 423 |

Note: Cells report regression coefficients, with standard errors (clustered at compound level) in parentheses. *, **, *** indicate significance at the 10, 5 and 1 percent level

Below is the regression output for the regressions on individual SanQoL attributes, rather than combined index values. Further below are the results of similar regressions but including interaction terms.

*Table D-3: Regression output underlying Table 4 results*

| | SanQoL attributes | | | | |
|---|---|---|---|---|---|
| | Disgust | Health | Shame | Safety | Privacy |
| Intervention toilet | 0.75***<br>(0.11) | 0.96***<br>(0.11) | 0.80***<br>(0.11) | 1.36***<br>(0.10) | 1.25***<br>(0.10) |
| Aged 60+ | -0.07<br>(0.17) | -0.07<br>(0.13) | 0.03<br>(0.15) | 0.06<br>(0.13) | -0.10<br>(0.12) |
| Female | -0.04<br>(0.09) | 0.08<br>(0.09) | 0.07<br>(0.10) | -0.30***<br>(0.08) | 0.02<br>(0.08) |
| Wealth index score | -0.10**<br>(0.05) | -0.03<br>(0.05) | 0.07<br>(0.05) | -0.00<br>(0.04) | 0.03<br>(0.04) |
| Constant | 1.62***<br>(0.10) | 1.38***<br>(0.10) | 1.55***<br>(0.10) | 1.43***<br>(0.10) | 1.59***<br>(0.10) |
| Observations | 423 | 423 | 423 | 423 | 423 |

Note: Cells report regression coefficients, with standard errors (clustered at compound level) in parentheses. *, **, *** indicate significance at the 10, 5 and 1 percent level. Attribute scores are on a 0-3 scale.

*Table D-4: Regression output underlying Table 4 interactions*

| | SanQoL attributes: gender interactions | | | | | SanQoL attributes: age interactions | | | | |
|---|---|---|---|---|---|---|---|---|---|---|
| | Disgust | Health | Shame | Safety | Privacy | Disgust | Health | Shame | Safety | Privacy |
| Intervention toilet | 0.69*** | 0.96*** | 0.66*** | 1.27*** | 1.27*** | 0.71*** | 0.92*** | 0.78*** | 1.42*** | 1.24*** |
| | (0.15) | (0.15) | (0.16) | (0.13) | (0.13) | (0.12) | (0.12) | (0.12) | (0.11) | (0.10) |
| Aged 60+ | -0.06 | -0.07 | 0.05 | 0.07 | -0.10 | -0.22 | -0.26 | -0.05 | 0.34 | -0.14 |
| | (0.17) | (0.14) | (0.16) | (0.13) | (0.13) | (0.28) | (0.24) | (0.28) | (0.24) | (0.22) |
| Female | -0.10 | 0.08 | -0.06 | -0.39*** | 0.04 | -0.03 | 0.09 | 0.08 | -0.31*** | 0.02 |
| | (0.15) | (0.16) | (0.16) | (0.14) | (0.14) | (0.10) | (0.10) | (0.10) | (0.08) | (0.08) |
| Wealth index score | -0.10** | -0.03 | 0.07 | -0.00 | 0.03 | -0.10** | -0.04 | 0.07 | -0.00 | 0.03 |
| | (0.05) | (0.05) | (0.05) | (0.04) | (0.04) | (0.05) | (0.05) | (0.05) | (0.04) | (0.04) |
| Intervention toilet # female | 0.11 | 0.00 | 0.26 | 0.18 | -0.03 | | | | | |
| | (0.20) | (0.19) | (0.20) | (0.17) | (0.16) | | | | | |
| Intervention toilet # over-60 | | | | | | 0.26 | 0.33 | 0.13 | -0.49* | 0.07 |
| | | | | | | (0.34) | (0.29) | (0.34) | (0.27) | (0.27) |
| Constant | 1.65*** | 1.38*** | 1.62*** | 1.48*** | 1.58*** | 1.64*** | 1.39*** | 1.56*** | 1.41*** | 1.59*** |
| | (0.12) | (0.12) | (0.12) | (0.11) | (0.11) | (0.10) | (0.10) | (0.10) | (0.10) | (0.10) |
| Observations | 423 | 423 | 423 | 423 | 423 | 423 | 423 | 423 | 423 | 423 |

Note: Cells report regression coefficients, with standard errors (clustered at compound level) in parentheses. *, **, *** indicate significance at the 10, 5 and 1 percent level. Attribute scores are on a 0-3 scale.

### E. Robustness checks

*Table E-1: Robustness checks for SanQoL and VAS*

| | **Outcome: SanQoL index value (0-1 scale)** | | | | | **Outcome: Sanitation VAS (0-10 scale)** | | | | |
|---|---|---|---|---|---|---|---|---|---|---|
| | (1) Headline GLMM (main model) | (2) Main model GEE | (3) Main model OLS | (4) GLMM with only 10% level covariates | (5) GLMM with theory-based covariates | (6) Headline GLMM (main model) | (7) Main model GEE | (8) Main model OLS | (9) GLMM with only 10% level covariates | (10) GLMM w/ theory-based covariates |
| Pour-flush toilet (Intervention) | 0.34*** (0.02) | 0.34*** (0.02) | 0.34*** (0.02) | 0.34*** (0.02) | 0.34*** (0.02) | 2.91*** (0.24) | 2.91*** (0.24) | 2.87*** (0.25) | 2.86*** (0.24) | 2.87*** (0.24) |
| Aged 60+ | -0.01 (0.03) | -0.01 (0.03) | -0.02 (0.03) | | -0.01 (0.03) | -0.15 (0.27) | -0.15 (0.28) | -0.36 (0.28) | | -0.13 (0.27) |
| Female | -0.01 (0.02) | -0.01 (0.02) | -0.01 (0.02) | | -0.01 (0.02) | -0.30* (0.16) | -0.30* (0.16) | -0.26 (0.17) | | -0.32** (0.16) |
| Wealth index score | -0.01 (0.01) | -0.01 (0.01) | -0.00 (0.01) | -0.01 (0.01) | | -0.07 (0.10) | -0.07 (0.10) | -0.09 (0.11) | -0.10 (0.10) | |
| Participant age (categorised) | | | | -0.00 (0.00) | | | | | -0.01 (0.03) | |
| Completed secondary school or above | | | | 0.05 (0.03) | | | | | 0.61** (0.25) | |
| Number of people sharing toilet stance | | | | | 0.00 (0.00) | | | | | 0.03* (0.02) |
| Shares toilet with other households | | | | | -0.14*** (0.03) | | | | | -1.14*** (0.34) |
| Renter | | | | | 0.03 (0.02) | | | | | 0.06 (0.20) |
| Constant | 0.50*** (0.02) | 0.50*** (0.02) | 0.50*** (0.02) | 0.49*** (0.02) | 0.58*** (0.04) | 4.28*** (0.20) | 4.28*** (0.19) | 4.30*** (0.20) | 4.08*** (0.23) | 4.89*** (0.38) |
| Observations | 423 | 423 | 423 | 423 | 424 | 423 | 423 | 423 | 423 | 424 |

Note: standard errors are shown in parentheses, which are clustered at the compound level; *, **, *** indicate significance at the 10, 5 and 1 percent level. In the main paper, Table 2 presents the mean of respondent age as a continuous variable. We categorised age in the publicly available replication dataset to maintain full anonymity, since several ages were shared by 5 people or fewer. We apply the categorised variable in these robustness checks for full replicability, but applying the continuous variable makes no difference to results.

*Table E-2: Robustness checks for WHO-5*

| | **Outcome: WHO-5 index** | | | | |
|---|---|---|---|---|---|
| | (1) Main model MEGLM (same as Table 4) | (2) Main model GEE | (3) Main model OLS | (4) MEGLM with only 10% level covariates | (5) MEGLM with theory-based covariates |
| Pour-flush toilet (Intervention) | 6.25**<br>(3.05) | 6.27**<br>(3.04) | 4.83<br>(3.15) | 6.56**<br>(3.02) | 6.21**<br>(2.88) |
| Aged 60+ | -12.95***<br>(2.91) | -12.87***<br>(3.30) | -15.98***<br>(3.41) | | -6.19**<br>(3.13) |
| Female | -3.31*<br>(1.94) | -3.34*<br>(1.88) | -2.19<br>(2.06) | | -1.29<br>(2.01) |
| Wealth index score | 0.98<br>(1.12) | 0.99<br>(1.19) | 0.78<br>(1.36) | 0.77<br>(1.14) | |
| Participant age (categorised) | | | | -1.67***<br>(0.37) | |
| Completed secondary school or above | | | | 1.44<br>(3.60) | |
| Has partner | | | | | -1.95<br>(2.09) |
| Pain scale | | | | | 7.23***<br>(2.24) |
| Problems walking scale | | | | | 5.79**<br>(2.71) |
| Constant | 57.01***<br>(2.40) | 57.00***<br>(2.39) | 57.39***<br>(2.47) | 60.68***<br>(2.76) | -4.87<br>(11.32) |
| Observations | 422 | 422 | 422 | 422 | 423 |

Note: standard errors are shown in parentheses, which are clustered at the compound level; *, **, *** indicate significance at the 10, 5 and 1 percent level

*Table E-3: Robustness checks for attribute level models (coefficients are odds ratios)*

| | Meologit for main attribute-level model | | | | |
|---|---|---|---|---|---|
| | Disgust | Health | Shame | Safety | Privacy |
| Intervention toilet | 5.58*** | 7.94*** | 5.51*** | 26.75*** | 65.66*** |
| | (1.70) | (2.52) | (1.46) | (11.15) | (41.82) |
| Aged 60+ | 0.93 | 0.78 | 1.08 | 1.08 | 0.83 |
| | (0.34) | (0.24) | (0.36) | (0.39) | (0.30) |
| female | 0.94 | 1.22 | 1.09 | 0.45*** | 1.07 |
| | (0.19) | (0.25) | (0.22) | (0.10) | (0.26) |
| wealth index score | 0.80* | 0.91 | 1.16 | 0.99 | 1.08 |
| | (0.10) | (0.11) | (0.12) | (0.11) | (0.14) |
| Observations | 423 | 423 | 423 | 423 | 423 |

Note: coefficients are odds ratios. Standard errors are shown in parentheses, which are clustered at the compound level; *, **, *** indicate significance at the 10, 5 and 1 percent level

*Table E-4: Robustness checks for attribute level interactions (coefficients are odds ratios)*

| | Meologit for attribute-level gender interaction | | | | | Meologit for attribute-level age interaction | | | | |
|---|---|---|---|---|---|---|---|---|---|---|
| | Disgust | Health | Shame | Safety | Privacy | Disgust | Health | Shame | Safety | Privacy |
| Intervention toilet | 4.87*** | 7.58*** | 4.42*** | 29.95*** | 71.50*** | 5.28*** | 7.36*** | 5.33*** | 35.60*** | 57.12*** |
| | (1.83) | (2.97) | (1.52) | (16.75) | (50.93) | (1.69) | (2.42) | (1.44) | (16.98) | (36.37) |
| Aged 60+ | 0.95 | 0.79 | 1.11 | 1.06 | 0.82 | 0.75 | 0.57 | 0.95 | 2.11 | 0.68 |
| | (0.36) | (0.25) | (0.37) | (0.39) | (0.31) | (0.41) | (0.28) | (0.47) | (1.02) | (0.28) |
| Female | 0.83 | 1.17 | 0.90 | 0.48*** | 1.12 | 0.96 | 1.24 | 1.10 | 0.41*** | 1.09 |
| | (0.24) | (0.36) | (0.23) | (0.13) | (0.31) | (0.20) | (0.26) | (0.22) | (0.10) | (0.26) |
| Wealth index score | 0.80* | 0.91 | 1.16 | 0.99 | 1.08 | 0.80* | 0.91 | 1.16 | 0.98 | 1.09 |
| | (0.10) | (0.11) | (0.12) | (0.11) | (0.14) | (0.10) | (0.11) | (0.12) | (0.11) | (0.14) |
| Female*Intervention interaction | 1.29 | 1.09 | 1.54 | 0.84 | 0.86 | | | | | |
| | (0.54) | (0.46) | (0.63) | (0.39) | (0.49) | | | | | |
| Age*Intervention interaction | | | | | | 1.49 | 1.74 | 1.29 | 0.26* | 2.06 |
| | | | | | | (1.12) | (1.11) | (0.89) | (0.18) | (2.22) |
| Observations | 423 | 423 | 423 | 423 | 423 | 423 | 423 | 423 | 423 | 423 |

Note: coefficients are odds ratios. Standard errors are shown in parentheses, which are clustered at the compound level; *, **, *** indicate significance at the 10, 5 and 1 percent level

### F. The role of sharing toilets

During study design it was anticipated that most people in our sample would be sharing toilets with other households, since this was a condition of MapSan enrolment four years previously. In the event, 90% of control and 88% of intervention households used shared toilets (Table 2 in manuscript). The households using private toilets were all single-household compounds, likely due to empty dwellings (driven by rental markets or migration) or changes in compound living arrangements since the intervention.

After discussing means by sharing status, we explore sharing in another set of regressions. In the set of robustness checks including all covariates hypothesised *ex ante* as influencing SanQoL, the binary covariate for sharing the toilet with other households was significant at the 1% level with a negative coefficient. This is explained by differences within the control group, where people using private PLs had substantially higher SanQoL than people using shared PLs (Figure F-1a). In the intervention group, sharing made little difference.

*Figure F-1: Differences between groups using private and shared toilets*

(a) SanQoL – mean index value by sharing and treatment group

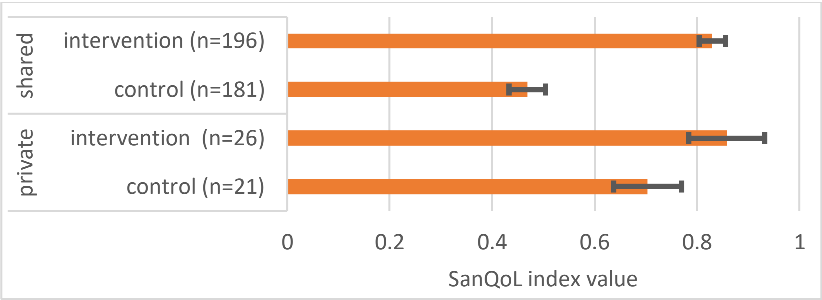


(b) VAS – mean score by sharing and treatment group

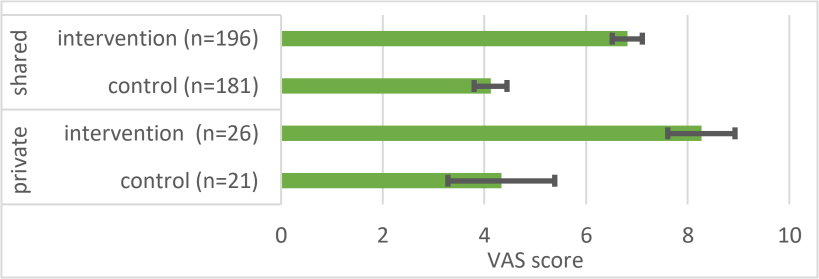


(c) WHO-5 – mean score by sharing and treatment group

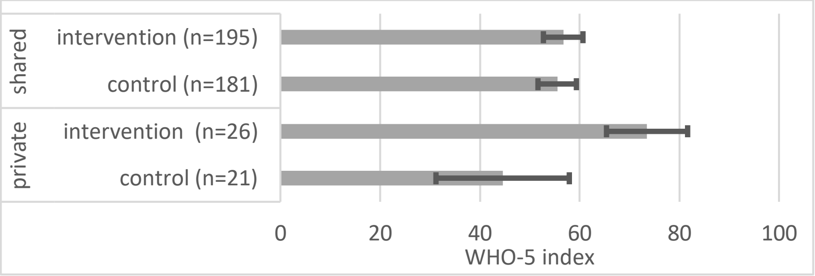


Note. error bars are 95% CIs

Table 2 in the manuscript demonstrated that the mean number of people per stance (cubicle) did not differ at the 5% level between intervention and control groups. This was also the case for analyses within private and shared, with 7.2 and 7.3 people on average using private toilets in control and intervention groups respectively, compared to 12.3 and 13.3 for shared toilets.

Considering VAS scores, a slightly different pattern was observed. Mean scores amongst people sharing intervention toilets were slightly lower than those not sharing, while there was no difference at the 5% level amongst controls (Figure F-1b). For both outcomes however, the intervention was associated with a substantial difference regardless of sharing status. For mental wellbeing, the picture is different again (Figure F-1c). There is a difference in WHO-5 amongst people using private toilets, but not amongst people sharing.

We ran a regression specified as per the headline results, but including a factorial interaction term between the intervention and the binary sharing variable (Table F-1). The results are easier to interpret in the light of Figure F-1. We make three interpretations from these results. First, amongst people sharing toilets with other households, there was strong evidence ($p<0.001$) that the intervention was associated with a difference in SanQoL of 0.37 (95% CI: 0.32 – 0.41). This is greater than the difference of 0.34 in the sample as a whole, and substantially larger than the difference of 0.15 (95% CI: 0.06 – 0.24) amongst those using private toilets. However, this is likely driven by the fact that SanQoL was already higher in the control group amongst those using private toilets (Figure F-1a) so scores didn't "have as far to travel" on a 0-1 scale.

Second, the opposite trend was seen in VAS scores. Amongst users of shared toilets the intervention was associated with a difference of 2.7 (95% CI: 2.2 – 3.2), while in private toilets it was associated with a difference of 3.9 (95% CI: 2.8 – 5.1). Scores in the intervention group simply increased more for private than shared (Figure F-1b). Third, for WHO-5 scores, there was no evidence of a difference amongst users of shared toilets (95% CI: -3.8 – 9.1), compared to a substantial difference of 27.9 amongst users of private toilets (95% CI: 13.0 – 42.8). This slightly odd result is again likely driven by the fact that WHO-5 scores were lower for households in the control group using private latrines (Figure F-1c). Note that comparisons between users of private toilets are based on a sample of only 20-25 people per treatment group.

Overall, the "private toilet" sub-group represents only 11% of the sample, and the intervention aimed to deliver high-quality shared sanitation rather than private sanitation. This invites the conclusion that the main results should be the focus. However, it is important that only four years after the intervention, the benefits of toilets which were meant to be shared were in fact being enjoyed by only one household (with mean size 7.3) in 12% of intervention compounds. To explore the quality of life effects of shared sanitation by comparison to private toilets, future studies would need to be adequately powered for this analysis.

*Table F-1: Interactions with sharing toilets*

| | SanQoL index value | VAS score | WHO-5 index score |
|---|---|---|---|
| Intervention | 0.15***<br>(0.05) | 3.94***<br>(0.59) | 27.91***<br>(7.59) |
| Aged 60+ | -0.02<br>(0.03) | -0.16<br>(0.27) | -12.73***<br>(2.89) |
| Female | -0.01<br>(0.02) | -0.33**<br>(0.16) | -3.61*<br>(1.93) |
| Wealth index score | -0.01<br>(0.01) | -0.10<br>(0.10) | 0.66<br>(1.12) |
| Shares toilet with other households | -0.24***<br>(0.04) | -0.27<br>(0.52) | 7.88<br>(6.65) |
| Intervention # shares toilet | 0.22***<br>(0.05) | -1.20*<br>(0.64) | -25.29***<br>(8.34) |
| Constant | 0.71***<br>(0.03) | 4.54***<br>(0.50) | 50.30***<br>(6.38) |
| Observations | 423 | 423 | 422 |

#### G. STROBE checklist

We report below which sub-sections provide information required in the STROBE checklist, using the form available at https://www.strobe-statement.org/checklists/

| | Item No | Recommendation |
|---|---|---|
| **Title and abstract** | 1 | (*a*) Indicate the study's design with a commonly used term in the title or the abstract<br><br>***Authors: the abstract notes that it is an observational comparison study of clusters previously enrolled in a non-randomised controlled trial*** |
| | | (*b*) Provide in the abstract an informative and balanced summary of what was done and what was found<br><br>***Authors: the abstract provides succinct background, methods that articulate the study design and outcomes, results, and their interpretation.*** |
| **Introduction** | | |
| Background/rationale | 2 | Explain the scientific background and rationale for the investigation being reported<br><br>***Authors: background and rationale are reported in the introduction*** |
| Objectives | 3 | State specific objectives, including any prespecified hypotheses<br><br>***Authors: the aim is stated in the last sentence of the introduction. Hypotheses are summarised in the "statistical analyses" sub-section of methods, with further rationale for them provided in Online Appendix B.*** |
| **Methods** | | |
| Study design | 4 | Present key elements of study design early in the paper<br><br>***Authors: there is a "study design" sub-section at the beginning of methods*** |
| Setting | 5 | Describe the setting, locations, and relevant dates, including periods of recruitment, exposure, follow-up, and data collection<br><br>***Authors: there is a specific "setting" sub-section in methods, including locations, and a map provided in Online Appendix A. The remaining aspects are reported in other parts of the methods section.*** |
| Participants | 6 | (*a*) *Cohort study*—Give the eligibility criteria, and the sources and methods of selection of participants. Describe methods of follow-up<br>*Case-control study*—Give the eligibility criteria, and the sources and methods of case ascertainment and control selection. Give the rationale for the choice of cases and controls<br>*Cross-sectional study*—Give the eligibility criteria, and the sources and methods of selection of participants<br><br>***Authors: the eligibility criteria are listed in the "participants" sub-section of methods, as are methods for sampling of participants, with additional detail provided in Online Appendix B.*** |
| | | (*b*) *Cohort study*—For matched studies, give matching criteria and number of exposed and unexposed<br>*Case-control study*—For matched studies, give matching criteria and the number of controls per case |

| | | |
|---|---|---|
| | | ***Authors: our study design did not use matching directly, but relied on the methods for identification of intervention/control compounds applied by the MapSan trial which are reported in the "study design" sub-section of methods.*** |
| Variables | 7 | Clearly define all outcomes, exposures, predictors, potential confounders, and effect modifiers. Give diagnostic criteria, if applicable<br><br>***Authors: Outcomes are defined in an "outcomes" sub-section of methods, with additional information provided in Online Appendix B. Other variables are defined in the "statistical analysis" section, with more detail in the data dictionary on LSHTM Data Compass*** |
| Data sources/ measurement | 8* | For each variable of interest, give sources of data and details of methods of assessment (measurement). Describe comparability of assessment methods if there is more than one group<br><br>***Authors: all data are primary and measured by household survey (see #7 above). Further detail on the administration of the survey are in the "participants" sub-section of methods, with further detail in Online Appendix B.*** |
| Bias | 9 | Describe any efforts to address potential sources of bias<br><br>***Authors: Efforts to address bias are described in the "participants" sub-section of methods, and also in the "study design" sub-section. Robustness checks to explore risk of bias in the analytical approach are described in the "sensitivity analysis" sub-section.*** |
| Study size | 10 | Explain how the study size was arrived at<br><br>***Authors: the sample size calculation is described in the "statistical analyses" section.*** |
| Quantitative variables | 11 | Explain how quantitative variables were handled in the analyses. If applicable, describe which groupings were chosen and why<br><br>***Authors: regression models are described in the "statistical analyses" section.*** |
| Statistical methods | 12 | (*a*) Describe all statistical methods, including those used to control for confounding<br><br>***Authors: covariates controlled for are described in the "statistical analyses" section.*** |
| | | (*b*) Describe any methods used to examine subgroups and interactions<br><br>***Authors: approaches to interactions with gender and being elderly are described as hypotheses in the "statistical analyses" section.*** |
| | | (*c*) Explain how missing data were addressed<br><br>***Authors: We state in the "statistical analyses" section that only 2 participants had missing data for outcomes or covariates (one for WHO-5, one for the wealth index). With 424 respondents each represents 0.2% of the sample and is not an important source of bias.*** |
| | | (*d*) *Cohort study*—If applicable, explain how loss to follow-up was addressed<br>*Case-control study*—If applicable, explain how matching of cases and controls was addressed<br>*Cross-sectional study*—If applicable, describe analytical methods taking account of sampling strategy |

| | | |
|---|---|---|
| | | ***Authors: we explain the approach to accounting for clustering in the "statistical analyses" section*** |
| | | (e) Describe any sensitivity analyses<br><br>***Authors: robustness checks are described in the "sensitivity analysis" sub-section*** |
| **Results** | | |
| Participants | 13* | (a) Report numbers of individuals at each stage of study—eg numbers potentially eligible, examined for eligibility, confirmed eligible, included in the study, completing follow-up, and analysed<br><br>***Authors: numbers of compounds are described in the participant flow diagram (Figure 1)*** |
| | | (b) Give reasons for non-participation at each stage<br><br>***Authors: reasons are explained in Figure 1*** |
| | | (c) Consider use of a flow diagram<br><br>***Authors: See Figure 1*** |
| Descriptive data | 14* | (a) Give characteristics of study participants (eg demographic, clinical, social) and information on exposures and potential confounders<br><br>***Authors: Participant characteristics are summarised in Table 2, separately for intervention and control groups.*** |
| | | (b) Indicate number of participants with missing data for each variable of interest<br><br>***Authors: the note to Table 2 states that one participant had missing data for the wealth index.*** |
| | | (c) *Cohort study*—Summarise follow-up time (eg, average and total amount)<br><br>***Authors: n/a*** |
| Outcome data | 15* | *Cohort study*—Report numbers of outcome events or summary measures over time<br><br>***Authors: n/a*** |
| | | *Case-control study*—Report numbers in each exposure category, or summary measures of exposure<br><br>***Authors: n/a*** |
| | | *Cross-sectional study*—Report numbers of outcome events or summary measures<br><br>***Authors: Distributions of outcomes are presented in histograms in Online Appendix C, separately for intervention and control groups, as are distributions of individual SanQoL attributes*** |
| Main results | 16 | (*a*) Give unadjusted estimates and, if applicable, confounder-adjusted estimates and their precision (eg, 95% confidence interval). Make clear which confounders were adjusted for and why they were included<br><br>***Authors: unadjusted and adjusted estimates with 95% CIs are provided in Tables 3 and 4. The rationale for confounder adjustment is provided in the "statistical analyses" section of methods.*** |
| | | (*b*) Report category boundaries when continuous variables were categorized |

| | | |
|---|---|---|
| | | ***Authors: the note to Table 2 states that in the replication dataset available online, we categorised age, household size and children under 14 to maintain full anonymity, since several values were shared by 5 people or fewer.*** |
| | | (*c*) If relevant, consider translating estimates of relative risk into absolute risk for a meaningful time period<br><br>***Authors: n/a*** |
| Other analyses | 17 | Report other analyses done—eg analyses of subgroups and interactions, and sensitivity analyses<br><br>***Authors: interactions are reported in the results section, with full results tabulated in Online Appendix D. Sensitivity analysis findings are described in the results section, with full tables in Online Appendix E.*** |
| **Discussion** | | |
| Key results | 18 | Summarise key results with reference to study objectives<br><br>***Authors: Key results are summarised in the first paragraph of the discussion.*** |
| Limitations | 19 | Discuss limitations of the study, taking into account sources of potential bias or imprecision. Discuss both direction and magnitude of any potential bias<br><br>***Authors: Limitations are discussed in two paragraphs towards the end of the discussion, including sources and direction of potential bias.*** |
| Interpretation | 20 | Give a cautious overall interpretation of results considering objectives, limitations, multiplicity of analyses, results from similar studies, and other relevant evidence<br><br>***Authors: Interpretation is undertaken in the first paragraph of our discussion, with potential mechanisms discussed in the second paragraph.*** |
| Generalisability | 21 | Discuss the generalisability (external validity) of the study results<br><br>***Authors: generalisability is considered in the final paragraph of the discussion*** |
| **Other information** | | |
| Funding | 22 | Give the source of funding and the role of the funders for the present study and, if applicable, for the original study on which the present article is based<br><br>***Authors: This work was supported by the Economic and Social Research Council through a PhD studentship. The fieldwork was funded by the Bill & Melinda Gates Foundation. The funders had no role in the identification, design, conduct, or reporting of the analysis. The original funders of the MapSan trial on which our study is based were the United States Agency for International Development and the Bill & Melinda Gates Foundation.*** |

**Give information separately for cases and controls in case-control studies and, if applicable, for exposed and unexposed groups in cohort and cross-sectional studies.*

#### H. Reflexivity Statement and checklist

*Reflexivity statement (Table 1 from Morton et al. 2021)*[23]

| Area | Question | Response |
|---|---|---|
| **Study conceptual-isation** | 1. How does this study address local research and policy priorities? | The National Strategy for Urban Water and Sanitation in Mozambique has the objective of universal access to sanitation services by 2025, but in 2020 only 61% of the urban population had access to "basic" sanitation. Sanitation decision-makers at the Maputo City Council are interested means by which they can improve on-site sanitation in informal settlements where sewerage expansion is unrealistic in the medium-term. Understanding which interventions most improve people's quality of life will help identify interventions most likely to achieve sustained uptake at scale, and all research collaborators buy into the importance of this. |
| | 2. How were local researchers involved in study design? | ZA led the fieldwork in 2018 for the qualitative study which informed the development of the SanQoL measure, and undertook discussions of methods for the present study with IR at that time. ZA and IR had also collaborated on a separate household survey in 2018 in the same setting, which informed the approach to sampling and data collection in the present study. ZA and IR refined the methods for the present study, especially the SanQoL questions and other outcomes, through discussion, fieldworker training, cognitive interviews and piloting. RN contributed to study design by inputting into the protocol and data collection instruments. |
| **Research management** | 3. How has funding been used to support the local research team(s)? | This study was funded under the Maputo Sanitation trial (MapSan) programme of research (clinicaltrials.gov, NCT02362932). The associated funding covered salary costs of ZA and RN for several years, as well as other staff at the Instituto Nacional de Saúde (INS) and WE Consult. The team has supported an INS researcher to successfully apply for a funded PhD programme, and has supported three grant proposals led by early career researchers at INS. |
| **Data acquisition and analysis** | 4. How are research staff who conducted data collection acknowledged? | The four members of the fieldwork team are named in the acknowledgements. |
| | 5. How have members of the research partnership been provided with access to study data? | The data are available open access online. |
| | 6. How were data used to develop analytical skills within the partnership? | As required for a PhD thesis, the analysis was conducted independently by IR with only limited support from the supervisors GG and OC. |
| **Data interpretation** | 7. How have research partners collaborated in interpreting study data? | ZA and IR discussed emerging trends in data as it came in. ZA and IR also discussed interpretation of observations for some variables which informed data cleaning and analysis. ZA and RN inputted into iterations of the manuscript. |
| **Drafting and revising for** | 8. How were research partners supported to develop writing skills? | As part of the PhD thesis, the first draft of the manuscript was completed independently by IR. |

| | | |
|---|---|---|
| **intellectual content** | 9. How will research products be shared to address local needs? | This study will be published open access. IR undertook a scoping of the health economics policy community in Mozambique while based there during data collection, and will share results of the study with identified stakeholders. There is a dissemination plan for the broader body of research, which will include engagement of urban sanitation stakeholders in Maputo upon IR's next travel to Mozambique. |
| **Authorship** | 10. How is the leadership, contribution and ownership of this work by LMIC researchers recognised within the authorship? | ZA and RN contributed to study design and data interpretation as outlined in the answers to questions #2 and #7 above. The study was part of IR's PhD thesis and predominantly his work, so he has the first author position. Of the more senior members of the study team (RN, OC, JB, CO, GG), all co-authors agree that OC played the most substantial role in guiding the study's methods and interpretation and in supervising this part of IR's PhD thesis, which is why he has the last author position. |
| | 11. How have early career researchers across the partnership been included within the authorship team? | There are two early career researchers within the authorship team, ZA (based in Mozambique) and IR (based in the UK). |
| | 12. How has gender balance been addressed within the authorship? | Four authors are male (IR, JB, CO, and OC) and three female (GG, ZA, and RN). |
| **Training** | 13. How has the project contributed to training of LMIC researchers? | This specific study did not include a training component. However, under the broader MapSan programme of research, Mozambican researchers from INS received training on different research methods related to epidemiology and laboratory skills. |
| **Infrastructure** | 14. How has the project contributed to improvements in local infrastructure? | This specific study has not directly contributed to improvements in local infrastructure. |
| **Governance** | 15. What safeguarding procedures were used to protect local study participants and researchers? | The participant information sheet was approved by ethical review committees in Mozambique and the UK. It details procedures for confidentiality, withdrawal, and complaints. Fieldwork team members worked in pairs, keeping in contact with ZA by phone on their location. Their training included guidance on what to do in the event of an emergency. |

#### *Checklist (Table 2 from Morton et al. 2021)*

| Area | Question | Response |
|---|---|---|
| **Engagement** | Has the research team engaged constructively with the reflexivity statement? | Yes, all questions have been completed in the table above. |
| **Co-development** | Have the research partners co-developed the research study? | Yes, as illustrated in responses to questions 2, 3, 7 and 10 in the table above. |
| | Does the study address priority research questions for the LMIC partner(s)? | Yes, see answer to question 1 above. |
| **Authorship** | Is there a LMIC partner who is the first or last author? | No. |
| | If not, what is the explanation? | Reasons are set out in response to question 10 above (PhD thesis, supervision, and level of support to the specific study) |

| | | |
|---|---|---|
| | How have LMIC early career researchers been incorporated as authors? | Yes, see answer to question 11 above. |
| **Dissemination** | How are data shared with LMIC partners to address research needs? | Data are available open access online. |
| | Is there open access funding to improve publication dissemination? | Yes, one of the funders (BMGF) ensures open access for any studies carried out with this funding. |
